# Peripheral Predictors of Speech in Noise Deficits in a Population Cohort

**DOI:** 10.64898/2026.01.27.26344934

**Authors:** Christopher R. Cederroth

**Affiliations:** Laboratory of Translational Auditory Neuroscience, Department of Physiology and Pharmacology, Karolinska Institutet, Stockholm, Sweden

**Author notes:** Christopher R. Cederroth, Karolinska Institutet, Dpt. of Physiology and Pharmacology, Solnavägen 9, 171 77 Stockholm, Sweden.

## Abstract

**Background:** Age-related hearing loss is the leading sensory deficit in older adults, yet audiometric thresholds at conventional frequencies often poorly predict speech understanding. Two competing hypotheses have emerged: extended high-frequency (eHF) hearing loss beyond 8 kHz may unmask variance in speech performance, while cochlear synaptopathy, inferred via auditory brainstem response (ABR) wave I amplitude reduction, may degrade temporal coding independent of audiometry. This study aims to identify the best predictors of speech in noise deficits in the general population.

**Methods:** This is a cross-sectional analysis of 526 ears from 263 tinnitus-free adults (178 [67.7%] women and 85 men based on self-reported gender) enrolled in the Swedish Tinnitus Outreach Project, a population-based cohort study. Participants underwent air-conduction audiometry from 0.5 to 16 kHz, loudness discomfort levels, distortion product otoacoustic emissions, click-evoked ABR, and standardized speech-in-noise and word recognition testing in a clinical setting. Their associations with age and speech outcomes were evaluated using multivariable regression models.

**Results:** eHF pure-tone average was the most age-sensitive auditory metric, accounting for 64% of age-related variance, compared with 16% for PTA_4_ (the pure tone average of audiometric thresholds at 0.5, 1, 2, 4 kHz). eHF PTA thresholds (pure-tone average at 10-16 kHz) predicted speech-in-noise performance substantially better than PTA_4_, otoacoustic emissions, or ABR measures. ABR wave I amplitude did not significantly contribute to speech outcomes, even in high-reliability recordings (ICC = 0.96), whereas prolonged wave I latency was associated with poorer word recognition, particularly in older adults. In young adults, delayed wave I latency minimally affected word recognition, but in older adults an increase in latency from approximately 1.0 to 1.8 ms was associated with about a 27-percentage-point reduction in word recognition scores.

**Conclusions:** These findings identify eHF audiometry as a dominant clinical predictor of both cochlear aging and speech outcomes, and challenge the translational relevance of ABR-based synaptopathy markers in heterogeneous human populations, with implications for hearing assessment, clinical trials, and early intervention targeting in aging cohorts.

## Introduction

Hearing loss is typically defined as a loss of >20 dB hearing levels (HL) at the pure tone average of audiometric thresholds of 0.5, 1, 2, 4 kHz (PTA_4_) [1]. Based on this gold-standard, hearing loss affects over 1.5 billion people globally, with prevalence rising steeply after age 50. Between 1990 and 2019, global moderate-to-complete hearing loss increased 79.1%, from 225.3 million to 403.3 million, with projections suggesting 2.45 billion affected by 2050 [1]. Beyond auditory function, hearing loss has emerged as a potentially modifiable risk factor for cognitive decline and dementia [2]. Cross-sectional studies show strong associations of mild cognitive impairment evidencing impaired functional and anatomical connectivity once corrected for β-Amyloid deposition [3]. Population-based cohort studies reveal that hearing loss associates with incident cognitive impairment and dementia [4]. However, in systematic reviews and meta-analyses that have evidenced a strong relationship between cognitive impairment or dementia, 80% of the studies limited their auditory assessment to 4 kHz [5]. Despite its clinical importance, the PTA4 poorly predicts real-world communication ability and speech understanding in challenging environments [6]. In addition, clinical pure-tone audiometry (up to 8 kHz) explains limited variance in word recognition and speech-in-noise (SPiN) performance, leaving clinicians unable to accurately predict who will struggle most with communication [7]. As a consequence, hearing loss is often detected only once communication is impacted, leading to a delay of near 10 years until interventions (e.g., hearing aids) occur [8], although these could potentially prevent cognitive decline [9].

Difficulties in understanding speech may occur in presence of a normal audiogram. Early detection of SPiN deficits may thus contribute to earlier interventions for hearing and cognitive interventions [10]. Interestingly, recent work shows that eHF pure-tone averages (10–16 kHz) robustly predict word and phoneme recognition in challenging acoustic environments [11]. Since cochlear damage from aging, noise exposure or ototoxicity preferentially affects the cochlear base, where frequencies above 8 kHz are encoded [12], measurement of extended high-frequency (eHF) thresholds (> 8 kHz) may reveal clinically relevant cochlear pathology that conventional PTA misses. Recent studies have suggested a contribution of cochlear synaptopathy (CS) to “hidden hearing loss” and SPiN deficits [13–15]. CS derives from pre-clinical work showing that moderate noise exposure can selectively destroy high-threshold, low-spontaneous-rate auditory nerve fibers at the inner hair cell synapse without elevating auditory brainstem thresholds, albeit decreasing the auditory brainstem response (ABR) Wave 1 amplitude of the auditory brainstem response [16–18]. This synaptic pathology theoretically persists even after threshold recovery, potentially explaining deficits in speech understanding in noisy environments despite normal or near-normal quiet thresholds. Post-mortem histopathology of human temporal bones found cochlear nerve degeneration to be associated with poorer word recognition performance [19]. Likewise, in young healthy individuals with normal hearing thresholds up to 16 kHz, those with lower ABR Wave I amplitudes had poorer performance on word recognition in noise or with time compression and reverberation [20], a correlation supported by an independent study from Bramhall et al. [21]. However, the validity of the ABR Wave I amplitude as a biomarker for CS and/or SPiN has been debated [22, 23]. Thus, whether a reduction in Wave I amplitude may serve a noninvasive proxy of CS or SPiN in humans remains to be established.

While prior studies have examined eHF [11], DPOAE [24, 25], or ABR markers [21] in isolation, or in small [26, 27], highly selected samples [20], the present analysis is, to my knowledge, the first to directly address these two hypotheses against one another: eHF hearing loss versus ABR Wave I amplitude as an index of SPiN in a large, unselected general population cohort with comprehensive, quality-controlled audiometric and electrophysiological data. The analysis relies on data from the Swedish Tinnitus Outreach Project (STOP) but focuses on healthy adults without tinnitus in order to convey a clearer understanding of the relationship between these features and SPiN in absence of tinnitus. Here, eHF pure-tone audiometry (10–16 kHz), high-reliability ABR recordings with rigorous quality control, DPOAEs and speech recognition measures (word and phoneme recognition in noise) were investigated. Classical multivariate statistical modeling was used to assess the independent explanatory power of eHF thresholds vs ABR markers after accounting for conventional audiometry and demographic factors.

## Materials and Methods

### Ethics statement

The study was approved by the Regional Ethics Review Board in Stockholm (Dnr: 2016/1468-31/2); all participants provided written informed consent.

### Study Population and design

This report is a cross-sectional analysis of data collected at a single clinical assessment visit. Adult participants (>18 years old) were drawn from the Swedish Tinnitus Outreach Project (STOP) [28], a population-based cohort recruited 2016–2020 via the LifeGene national health study [29]. As a result, the cohort naturally includes a substantial proportion of tinnitus-free adults who volunteered as part of the broader population-based study. LifeGene invited all of their participants (N = 26,282) to join STOP, of whom 5,593 completed the STOP survey (2,588 reported no tinnitus, 1,131 occasional and 1,466 constant tinnitus, 403 were uncertain and 5 did not answer). Tinnitus-free participants (no tinnitus on any occasion, confirmed at the clinical session) were selected for this analysis to avoid confounding of ABR and audiometric measures by tinnitus-related central changes [28, 30]. The prevalence of tinnitus in the LifeGene cohort and the characterization of these participants has been previously reported [31–33]. Compared with tinnitus participants in STOP [28], the tinnitus-free subsample did not differ significantly in age (*p* = 0.21), gender ratio (*p* = 0.44), or conventional audiometric thresholds (PTA_4_, *p* = 0.19), suggesting they are broadly representative of the wider STOP population in the domains relevant to this study. Participants did not receive any financial compensation.

The participant flow diagram is reported in **Figure 1**. The present analysis included 263 participants (N = 526 ears) who (1) reported no tinnitus (neither occasional nor constant) (2) completed audiometry from 0.125–16 kHz, (3) completed speech-in-noise testing (N = 511 ears); click ABR was recorded in a subset of 168 participants (N = 336 ears). All were Swedish residents, and comprised 87% individuals with Swedish origin. Participants were highly educated (75% University level) and none were hearing aid or cochlear implant users. The ESIT screening questionnaire [34, 35] was added to the platform in November 2018 and was answered by a subset of participants (n = 32 missing). This allowed to determine that none were diagnosed for the following ear dysfunctions: ear barotrauma, Ménière’s Disease, Acoustic Neuroma, and Otosclerosis. Less than 2% of the participants had acoustic trauma, chronic otitis, or sudden hearing loss). Chemotherapy and head and neck radiotherapy was reported by 5 participants. Prevalence of other diseases is reported in **Table S1**.

**Figure 1.**
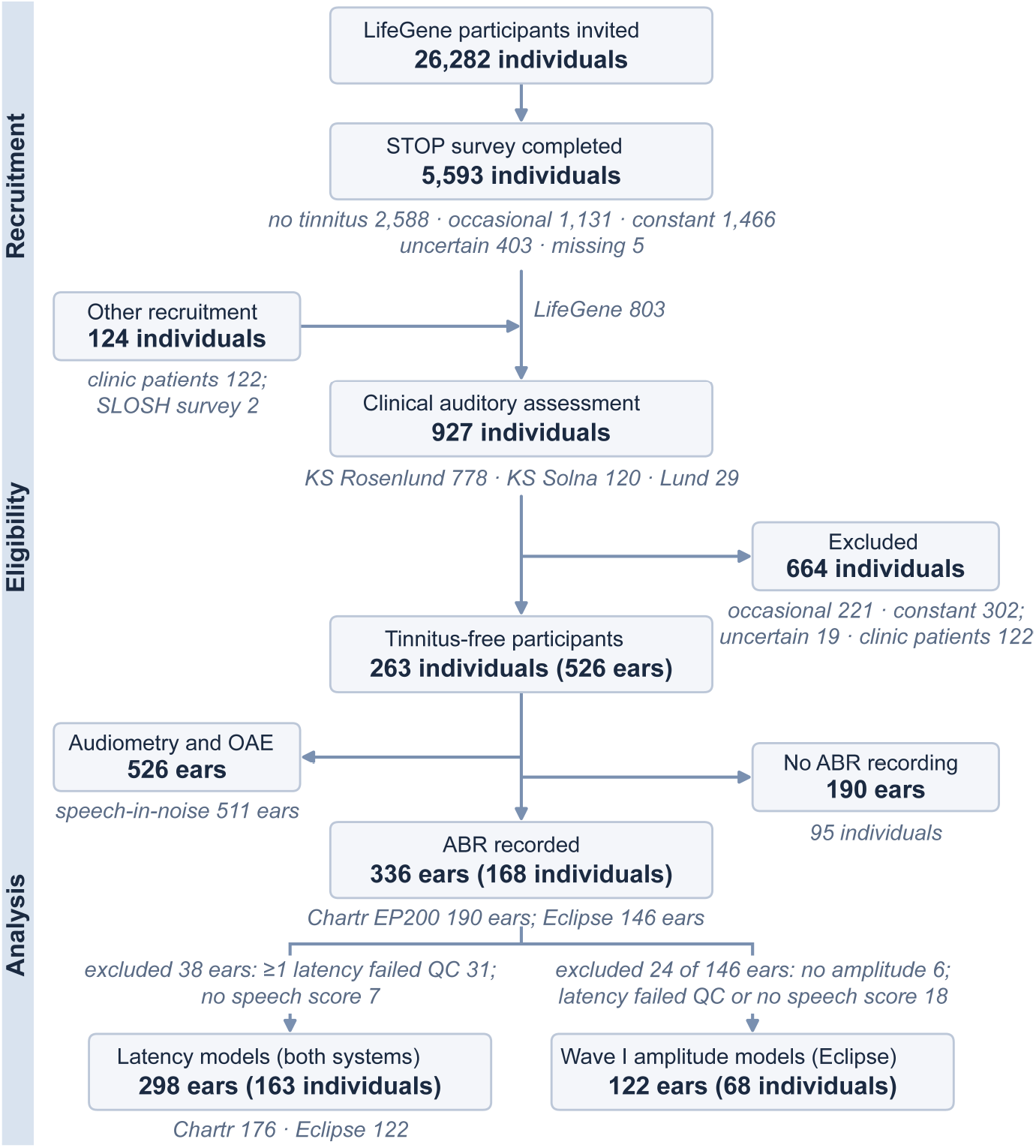
Participant flow diagram. Overview of participants excluded and recruited for the electrophysiological study. Of 26,282 LifeGene participants invited, 5,593 completed the STOP survey. Of 927 participants assessed clinically, 664 were excluded, leaving 263 tinnitus-free participants (526 ears). Of 168 participants with an ABR recording (336 ears), 298 ears from 163 participants had complete data for the latency models (both systems) and 122 ears from 68 participants for the Eclipse-only Wave I amplitude models. Univariate analyses used all ears available for each measure: audiometry and OAE, 526 ears; speech-in-noise, 511 ears; Wave I, III and V latency, 324, 322, 321 ears (wave-specific quality control); Eclipse Wave I amplitude, 140 ears. Each participant was recorded on a single ABR system.

Gender was self-reported in the STOP survey using item 2 of the Tinnitus Sample Case History Questionnaire (TSCHQ; response options male/female) for all 263 participants (178 women, 85 men) [36]. Sex assigned at birth was additionally recorded with item A2 of the ESIT-SQ [34] for 231 participants and agreed with self-reported gender in 226. Because the TSCHQ item does not distinguish gender identity from sex assigned at birth, analyses refer to self-reported gender, and no inference on sex-linked biological mechanisms is made. Study size was determined by the number of eligible tinnitus-free participants who completed the clinical assessment during the recruitment period. No a priori sample-size calculation was performed for the present analysis. Sex and Gender Equity in Research (SAGER) and Strengthening the Reporting of Observational Studies in Epidemiology (STROBE) guidelines were used to ensure clear, transparent and complete reporting [37, 38].

### Audiometry and otoacoustic emissions

Consenting tinnitus and non-tinnitus participants located in the Stockholm area or Skåne and who had answered large tinnitus survey [28] were invited for a battery of auditory measurements in a clinical setting. For external validity, auditory measurements were performed on 927 participants between August 2016 and December 2019 at three different sites: two different hospital audiology clinics in Stockholm (Karolinska Sjukhuset or KS Rosenlund, n = 778; and KS Solna, n = 120) and at the Department for Logopedics, Phoniatrics, and Audiology at Lund University (n = 29). For internal validity, the auditory measures and ABRs were collected by four licensed audiologists.

The session started with a short interview, confirming the participants’ tinnitus statuses, and otoscopy and tympanometry (Madsen OTOflex 100 tympanometer, Otometrics; #8-04-04900, GN Resound AB, Göteborg) was performed. The DPOAEs were measured using the Madsen Capella 2 (Madsen Capella 2, Otometrics; #8-04-15320, GN Resound AB, Göteborg) with L1 = 65 dB SPL and L2 = 55 dB SPL at ten points per octave between F2 = 1 kHz and F2 = 10 kHz. Pure tone audiometry was performed at standard (.125–8 kHz) and high (8–16 kHz) frequencies using a Madsen Astera 2 clinical audiometer (MADSEN Astera2, Otometrics; #8-04-13100, GN Resound AB, Göteborg) and HDA 200 headphones (Sennheiser) using fixed-frequency Bekesy audiometry with a pulsed pure tone (550 ms, 50% duty cycle). Bone conduction thresholds were verified manually for thresholds >20 dB HL between 0.125 and 4 kHz.

Speech-in-noise (SPiN) testing was performed using the clinical standard method and material in Sweden, presenting 50 CNC-words in +4dB SNR per ear at a level judged comfortable by the participants [39]. Participants repeated back isolated words (WS; 50 key words) and individual phonemes (PS; 3 phonemes per word, accuracy scored by consonant clusters). WS was quantified as the percentage of words in noise repeated correctly, and PS was estimated as percentage of phonemes in noise repeated correctly. Both measures were converted to percent correct (0–100%), where higher scores indicate better performance.

The LDLs were assessed after SIN at all threshold frequencies. A 1.5-second pure tone was presented at 60 dB HL, increased by 5 dB, and presented again. The subject was instructed to inform the tester if the tone presented would have been uncomfortable to listen to for a longer period (1–2 minutes). The session concluded with measurements of the ABR, detailed below.

### Auditory Brainstem Response (ABR)

Two setups for the ABR recordings were used in this study, the Madsen EP200 Chartr (ICS Chartr EP 200, Otometrics; #8-04-12701, GN Resound AB, Göteborg) and the Eclipse (Interacoustics, Stockholm) with ER-3A insert earphones and gold-wrapped tip trodes. The settings for both systems were identical, with high and low pass filters of .1 and 3 kHz, respectively, with 100 µs click stimuli of alternating polarity presented at 9.1 clicks/s at 90 dBnHL through insert earphones, with contralateral masking of −40 dB relative to the stimulus ear. Each recording consisted of 2,000 accepted clicks. The participants were relaxed in a reclined position in a dimly lit room during the recording. An audiologist identified relevant ABR features, i.e., wave I, III, and V peaks and troughs, then double-checked and, if necessary, corrected by two independent licensed audiologists blinded to the age of the participant. Amplitude was defined as peak to trough. In the present study, 190 non tinnitus ears were collected using the Chartr EP (Otometrics) and 146 ears with the Eclipse (Interacoustics). ABR Waves (N = 331, n = 5 missing) were classified as having good morphology (n = 151, 46%), fair morphology (n = 31, 9%), no clear wave forms for waves I (n = 2, 1%), III (n = 2, 1%), V (n = 4, 1%), and poor morphology (n = 141, 43%). In our previous study [28], test–retest reliability was established for these two devices: ICC for latencies 0.77–0.96 (good-to-excellent), ICC for amplitudes 0.26–0.47 (poor) on Chartr, 0.18-0.48 (poor) for Wave III and V but 0.96 (excellent) for Wave I amplitude on Eclipse.

### Statistical analysis

Age-related trajectories were modeled for all audiometric, OAE, speech, and ABR measures using separate polynomial regressions with age as the predictor and each variable [Pure-tone average at 0.5, 1, 2, and 4 kHz (PTA4), eHF pure-tone average at 10-16 kHz, DPOAE amplitude at 5,273 Hz, Word Score (WS), Phoneme Score (PS), Wave I amplitude, Wave I latency, Wave III latency, Wave V latency] as the outcome. For each measure, linear, quadratic, and cubic age models with age mean-centered were fitted, and the best functional form using hierarchical likelihood ratio tests (LRT) were selected in a sequential testing framework: linear models were compared to quadratic models, and if the quadratic term was significant (*p* < 0.05), quadratic models were compared to cubic models. The polynomial order was selected based on this stepwise LRT approach rather than AIC, to prioritize biological interpretability and avoid overfitting [40]. R^2^, LRT *p*-values comparing nested models, and the selected polynomial order for each relationship were reported. Wave I amplitude analyses were restricted to the Eclipse subset, whereas all other variables used the full cohort. The best-fitting age functions from these models were then used to generate the smooth age trajectories.

Univariate linear regression models were constructed with WS or PS as the outcome and audiometric (PTA4, eHF PTA, OAE), or ABR (Wave I amplitude, Wave I latency, Wave III latency, Wave V latency) predictors separately, using centered continuous predictors. Polynomial terms for each predictor were selected using the same hierarchical LRT approach (α = 0.05), testing linear vs. quadratic, then quadratic vs. cubic forms sequentially. Models were evaluated using R^2^, LRT p-values, and parameter estimates (β, SE, t, *p*), ensuring consistency in polynomial selection across univariate and multivariable frameworks [40]. Covariates were chosen a priori on subject-matter grounds rather than by data-driven variable selection: age and gender were treated as potential confounders, auditory measures were entered in prespecified blocks, and data-driven selection was limited to the polynomial order of each predictor. The Age x Gender interaction on WS and PS was examined in an exploratory model including age, age^2^ and gender. For the multivariable analysis predicting WS and PS, a prespecified four-phase hierarchical modeling strategy was followed, with each phase compared with the previous one using nested-model F tests:

- Phase 0 included demographic predictors only (age, age^2^, gender)
- Phase 1 added peripheral hearing-loss predictors (PTA4, EHF PTA, OAE) with LRT-selected polynomial terms
- Phase 2 added ABR latencies (Wave I, Wave III and Wave V) with LRT-selected polynomial terms to capture neural timing effects
- Phase 3 added the biologically motivated Age x Wave I latency interaction to test whether neural timing shows differential age-related vulnerability. Incremental variance explained (ΔR^2^) and nested-model F-test *p*-values were computed for each phase transition.

All analyses involving Wave I amplitude were restricted to the Eclipse subset (N = 140 ears; 134 with speech scores), where reliability is excellent (ICC = 0.96) [28]. Analyses of audiometry, OAE, and Wave latencies used the full cohort (N = 526 ears for audiometry, N = 511 ears for speech outcomes, N = 324 ears for Wave I latencies, N = 322 ears for Wave III latencies, N = 321 ears for Wave V latencies). The final latency models (Phase 3) were fitted to ears with complete data from both devices (n = 298 ears; Chartr, n = 176; Eclipse, n = 122) and did not include Wave I amplitude. Wave I amplitude was examined in a separate Eclipse-only sequence (n = 122 ears from 68 participants) in which Wave I amplitude (Phase 4) and the Age x Wave I amplitude interaction (Phase 5) were added to the Phase 3 model.

Multicollinearity was assessed via variance inflation factors (VIF < 5 for all models). Robust heteroscedasticity-consistent standard errors (HC3) from the sandwich–car framework were used because Breusch–Pagan tests indicated variance heterogeneity (studentized test for the final latency models: WS, χ²(17) = 35.04, p = 0.006; PS, χ²(17) = 43.49, p < 0.001), and HC3-based inferences are reported for all regression coefficients [41–43]. Model fit was assessed using adjusted R^2^, and diagnostic residual analyses in the complete-case subset (n = 298 ears), with sensitivity checks confirming that exclusion of extreme outliers did not materially change the pattern of significant predictors. Statistical significance was defined as α = 0.05 (two-tailed), and *p*-values are reported in scientific notation for values < 0.0001 in the main and supplementary tables.

### Unit of analysis, missing data and sensitivity analyses

The unit of analysis was the ear, and both ears of a participant were entered as separate observations when available. Because HC3 standard errors do not account for the correlation between the two ears of a participant (between-ear correlation for WS, r = 0.63), the final multivariable models were re-estimated with participant-clustered jackknife (CR3) robust standard errors with Satterthwaite small-sample degrees of freedom [44, 45]. A further sensitivity analysis added recording device as a covariate to the latency models. Missing data were not imputed: each analysis used all ears with non-missing values for the variables involved, and the final multivariable models were fitted to ears with complete data on all predictors and outcomes (complete-case subset, n = 298 ears from 163 participants, 135 of whom contributed both ears). Age groups for descriptive stratification were defined a priori in 10-year bands (<35, 35–44, 45– 54, 55–64, 65–74 and ≥75 years). For the WS–PS slope comparison, the complete-case subset was dichotomized at its median age (46 years).

All analyses were performed in R version 4.4.0 (R Foundation for Statistical Computing) using the tidyverse (v2.0.0) for data wrangling, car (v3.1-2) for regression diagnostics and HC3 standard errors, sandwich (v3.0-2) for robust variance estimation, clubSandwich for cluster-robust (CR3) inference, broom for model summaries, and ggplot2 (v4.0.0) with patchwork (v1.2.0) for figure making.

## Results

### Cohort characteristics

The STOP cohort comprised 526 ears from 263 participants aged 20 to 84 years (mean ± SD: 49.2 ± 13.6 years; **Table 1**), comprising 178 women (67.7%) and 85 men (32.3%). The study population was overall highly educated and with low disease prevalence (**Table S1**). Participants were measured in a clinical environment with a sound proof booth, with EC marked clinical devices. Overall, the cohort exhibited clinically normal hearing thresholds (cut-off 20 dB HL) in the conventional frequency range (PTA₄ 0.5–4 kHz: 5.2 ± 10.7 dB HL), which is the clinical gold-standard used by the WHO for classifying hearing impairment severity [1]. In contrast, substantial variability was found in eHF hearing loss (eHF PTA 10–16 kHz: 36.5 ± 22.5 dB HL, range: –12.2 to 78.0 dB HL). Loudness Discomfort Levels (LDLs) plateaued near ceiling values of the medical device (75.6–93.9 dB HL). Distortion Products of Otoacoustic Emissions (DPOAEs), a measure of outer hair cell (OHC) health and function, at 5.273 kHz averaged 19.8 ± 10.1 dB signal to noise ratio (SNR). Speech perception performance (ranging from 0 to 100% accuracy) was tested on 511 ears with hearing abilities in the standard speech weighted noise (+4 dB SNR) was robust, with mean word scores (WS) of 75.1 ± 10.3% and phoneme scores (PS) of 89.2 ± 6.0%. Auditory brainstem response measurements (available for 336 ears; **Table 1**, **Fig. 1**) were performed on two different medical devices: the Chartr EP (Otometrics) and the Eclipse (Interacoustics). In our former study, test-retest reliability was moderate (ICC>0.5) for all Wave latencies in both devices, but poor (ICC<0.5) for all Wave amplitudes, with the exception of the Eclipse Wave I amplitudes being excellent (>0.9) [28]. As a consequence, only Wave I amplitudes for the Eclipse are reported (0.4 ± 0.2 µV, n = 140 ears), and the Wave latencies (from both devices) were 1.4 ± 0.2 ms for Wave I, 3.6 ± 0.2 ms for Wave III, and 5.4 ± 0.4 ms for Wave V. The I-V interwave latency was 4.0 ± 0.3 ms (**Table 1**). Audiometry and OAEs were available for all 526 ears (263 participants), speech scores for 511 ears (257 participants), ABR latencies for 321–324 ears (167–168 participants) and Eclipse Wave I amplitude for 140 ears (72 participants); 298 ears from 163 participants had complete data for the final multivariable models (**Fig. 1**).

**Table 1.** Demographics from 263 tinnitus-free STOP participants (526 ears).

| Variable | Mean ± SD or N (%) | Range |
|---|---|---|
| Age (years) | 49.2 ± 13.6 | 20.0 to 84.0 |
| Gender, women (participants) | 178 (67.7%) | - |
| Gender, men (participants) | 85 (32.3%) | - |
| PTA4 (0.5-4 kHz) dB HL | 5.2 ± 10.7 | -7.4 to 100.0 |
| EHF PTA (10-16 kHz) dB HL | 36.5 ± 22.5 | -12.2 to 78.0 |
| OAE 5273 Hz (dB SNR) | 19.8 ± 10.1 | -31.0 to 44.6 |
| Word score (%) | 75.1 ± 10.3 | 12.0 to 94.0 |
| Phoneme score (%) | 89.2 ± 6.0 | 48.0 to 97.3 |
| Wave I amplitude (Eclipse uV) | 0.4 ± 0.2 | 0.1 to 1.0 |
| Wave I latency (ms) | 1.4 ± 0.2 | 1.0 to 1.9 |
| Wave III latency (ms) | 3.6 ± 0.2 | 3.0 to 4.2 |
| Wave V latency (ms) | 5.4 ± 0.4 | 4.5 to 6.3 |
| I–V inter-wave latency (ms) | 4.0 ± 0.3 | 3.2 to 4.8 |

Age and gender are reported per participant (n = 263); all other variables per ear. Ears available: PTA<sub>4</sub>, eHF PTA and OAE, n = 526; word and phoneme scores, n = 511; Wave I amplitude (Eclipse only), n = 140; Wave I, III and V latencies, n = 324, 322 and 321; I–V inter-wave latency, n = 312. Seven ears from six participants had PTA<sub>4</sub> > 40 dB HL (maximum 100 dB HL); none of them had complete ABR data, so they did not enter the multivariable models.

### Auditory performance declines with age

Age-stratified analyses revealed progressive deterioration across auditory domains (**Fig. 2a, Figure S1, Table S2**): eHF thresholds worsened from 13.8 ± 15.2 dB HL in participants <35 years to 69.0 ± 6.2 dB HL in those ≥75 years. In contrast, PTA_4_ remained within normal range according to WHO standards (cut-off = 20 dB HL), even in the older group (≥75 years: 18.5 ±11.3 dB HL). LDLs had near ceiling effects and tolerance to loud sounds at 8 kHz increased with age (**Fig. 2b**). Both PTA and LDLs showed device limitations in the eHF with maximal dB HL values achieved for 10 kHz (93 dB HL); 12.5 kHz (85 dB HL); 4 kHz (75 dB HL) and 16 kHz (55.5 dB HL). DPOAEs (hereafter referred to as OAEs), showed also an age-progressive decline, most evident for the 5.273 kHz, with a 24.5 ± 8.4 dB SNR in participants <35 years to 7.9 ± 10.1 dB SNR in those ≥75 years (**Fig. 2c**). WS and PS declined from 78.3 ± 6.5% to 55.6 ± 19.4%, and 90.9 ± 3.7% to 77.8 ± 12.3% respectively, across the same age ranges (**Fig. 2d,e**). Wave I amplitude (Eclipse only) demonstrated an age-related decline from 0.5 ± 0.2 µV in younger participants (<35 years) to 0.3 ± 0.1 µV in the 65–74 age group (**Fig. 2f**). Wave latency increased with age in Wave I (from 1.3 ± 0.1 ms in <35 years to 1.5 ± 0.2 ms in 65–74 years) and Wave V (from 5.3 ± 0.3 ms to 5.6 ± 0.3 ms; **Fig. 2g-i**), with corresponding prolongation of central conduction time (I–V inter-wave latency: 3.9 ± 0.3 ms to 4.1 ± 0.3 ms). The ABR data collected on two platforms (Chartr n = 190, Eclipse n = 146) showed systematic offset (0.21 ms latency, *p* <0.001) but no device and predictor interactions (all *p* > 0.10), indicating findings replicate across independent measurement systems. The mean and standard error of the mean (SEM) latency values obtained from the two devices using equal settings are reported for each age group (**Fig. 2g-i**). Stratification by gender and eHF tertiles is reported in **Table S3,4**.

**Figure 2:**
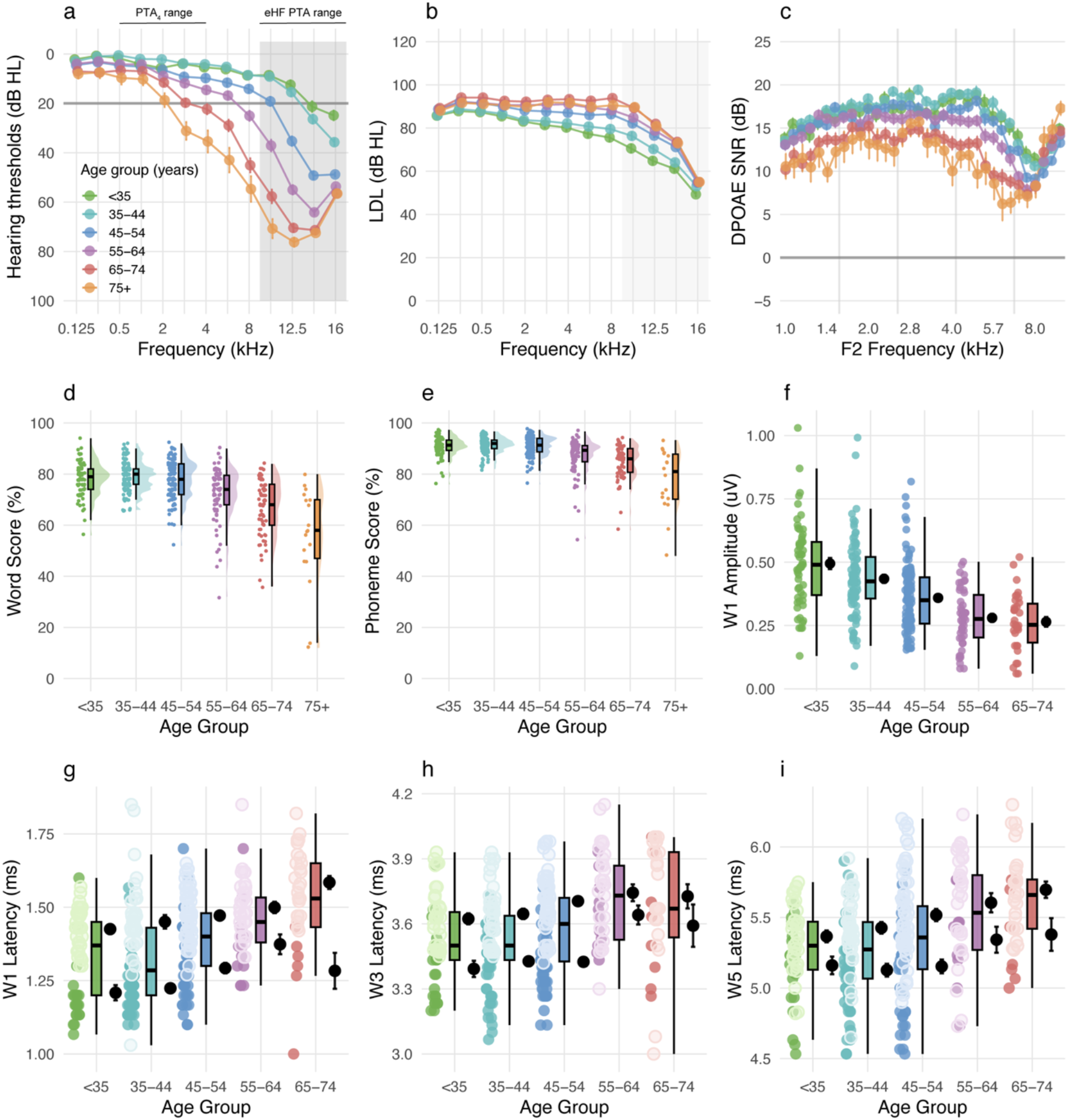
Audiometric patterns across age groups in the STOP cohort. Age-stratification of pure-tone audiometry (a), loudness discomfort levels (b), distortion products of otoacoustic emissions (c). Raincloud plots for word (d) and phoneme (e) scores individual points (left), summary statistics as median and IQR (middle) and representation of distribution with one-sided violin plots (right). Auditory brainstem responses are displayed for Wave I amplitude (f), and latencies of Wave I (g), III (h), V (i). Wave I amplitude are only shown for Eclipse hardware, the Chartr ones being unreliable. For Wave latencies, data from the Chartr and the Eclipse are distinguishable in the raw data (left) with lighter coloring for the Chartr, corresponding to different means and sem displayed on the right of the bar plot as “c” and “e” respectively. Age groups are displayed in different colors: <35 (green), 35-44 (light blue); 45-54 (blue); 55-64 (violet), 65-74 (red), and ≥75 (orange). Data relates to **Figure S1 and Tables S2-4**.

### Nonlinear Relationships Between Auditory Function and Aging

Guided by the non-linear, age-stratified patterns in **Fig. 2**, each audiometric and physiological variable were modeled as a function of age using univariate polynomial regression (linear vs. quadratic vs. cubic fits; **Fig. 3, Table S5**). Consistent with a former study from Bugannim et al. [6], PTA₄ showed a modest but significant quadratic increase with age (R^2^ = 0.158, quadratic *p* = 0.0057). In contrast, eHF PTA exhibited a much steeper age-related elevation with high overall fit (R² = 0.641) but no additional improvement from adding a quadratic term (quadratic p = 0.548), indicating an approximately linear worsening of eHF thresholds across the adult lifespan (**Fig. 3a,b**), corroborating former work from Motlagh Zadeh et al. [11]. OAE amplitude declined with age in a curvilinear fashion (R^2^ = 0.237, quadratic *p* = 0.009; **Fig. 3c**), while both WS and PS showed pronounced accelerating decline captured by strong quadratic components (WS: R^2^ = 0.296, quadratic *p* <0.0001; PS: R^2^ = 0.283, quadratic *p* <0.0001), confirming an inverted-U relationship published by Bugannim et al. [6], whereby speech perception performance initially improves with age before plateauing or declining (**Fig. 3d,e**). The Age^2^ coefficients were negative for both outcomes (WS: β = −0.013, *p* < 0.001; PS: β = −0.0077, *p* < 0.001), indicating that gains diminish progressively across the lifespan (**Table S6**). Gender exerted only marginal effects on speech perception. When age was included in the model, women performed modestly better than men (WS: +1.82 points, *p* = 0.034; PS: +1.00 point, *p* = 0.05), though the gender effect on phoneme scores was marginally significant. Critically, the Age x Gender interaction was not significant for either outcome (WS: *p* = 0.271; PS: *p* = 0.067), providing no clear evidence that age-related trajectories of speech perception differed between women and men (**Table S6**), although the study was not powered to detect such interactions. Among ABR measures, Wave I amplitude (Eclipse only) decreased with age but was adequately described by a linear fit (R^2^ = 0.195, quadratic *p* = 0.893, **Fig. 3f**), whereas Wave I, III and V latencies all displayed statistically significant quadratic age effects (Wave I: R^2^ = 0.153, p = 0.007; Wave III: R^2^ = 0.102, *p* = 0.048; Wave V: R² = 0.127, *p* = 0.028), consistent with subtly increased delay at older ages (**Fig. 3g–i, Table S5**). These polynomial fits confirm that several key predictors (PTA₄, OAE, WS/PS, and brainstem latencies) exhibit non-linear age relationships, providing the rationale for the subsequent univariate polynomial analyses of WS and PS and for including higher-order terms only where formally justified in the multivariable models.

**Figure 3:**
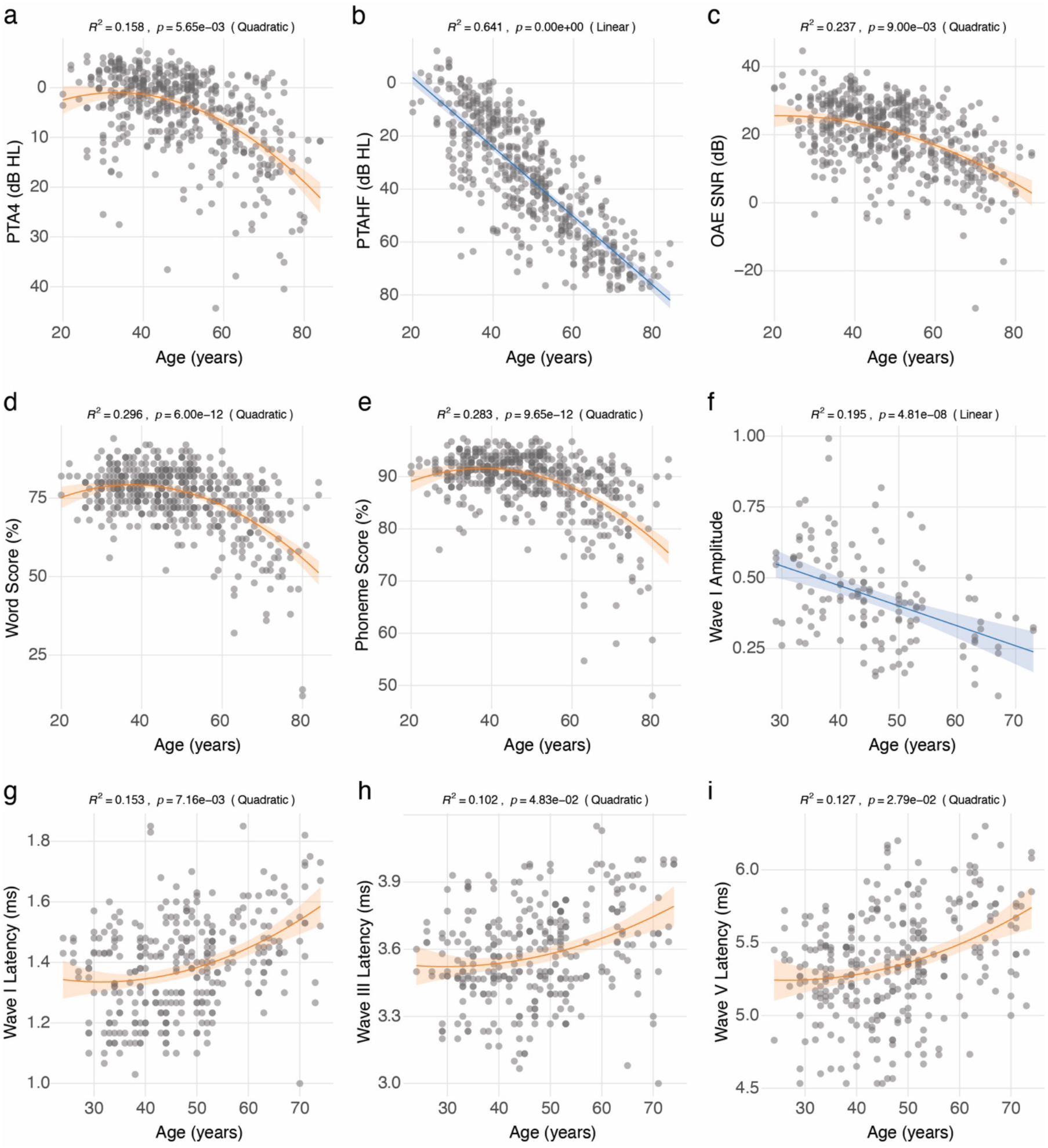
Age related changes in audiometric and electrophysiological variables. Univariate polynomial regressions (linear vs. quadratic vs. cubic fits) from Fig.2 are displayed for (a) PTA4 (average of 0.5, 1, 2, 4 kHz), (b) EHF PTA (average of 10, 12.5, 14, 16 kHz), (c) DPOAEs at 5273 Hz (OAE5273), (d) WS, (e) PS, (f) Wave I amplitude (Eclipse only), and latencies for Wave I (g), III (h) and V (i) in function of age. Best fits are reported for linear (blue), and quadratic (orange) with shaded regions as 95% confidence intervals, and R^2^ and *p* values on top of the respective panels. Data relates to **Table S5**.

### Influence of audiometric measures on speech perception

To directly answer the primary research question and identify which peripheral measures best predict SPiN outcomes in this population cohort, a hierarchical multivariable regression model was constructed, informed on the preceding univariate analyses (**Figures 2–3**) that established the age-related trajectories of each measure and their bivariate relationships with speech outcomes, ultimately providing the justification for predictor selection and polynomial terms in the final models. To optimize model specification, whether audiometric and physiological predictors exhibited nonlinear relationships with speech perception outcomes was systematically evaluated using likelihood ratio testing (LRT), fitting univariate polynomial models (linear, quadratic, cubic) for each predictor against word score (WS) and phoneme score (PS). EHF pure-tone average (eHF PTA, 10–16 kHz) was the single strongest peripheral predictor of speech performance (**Fig. 4, Table S7**): cubic models explained substantially more variance in WS than conventional PTA₄ (R^2^ = 0.345 vs. 0.221; cubic *p* = 0.0014 for eHF PTA, *p* = 0.047 for PTA₄; **Fig. 4a,b**), with an even larger advantage for PS (eHF PTA R^2^ = 0.361 vs. PTA₄ R^2^ = 0.247; eHF PTA cubic *p* = 0.0001, PTA₄ quadratic *p* <0.0001; **Fig. 4g,h**). Both audiometric measures showed non-linear relationships with speech outcomes (cubic for eHF PTA; cubic for PTA₄ with WS and quadratic with PS), consistent with non-linear effects of hearing loss on speech perception rather than a simple monotonic decline. Critically, eHF PTA outperformed PTA₄ by 12.4 percentage points for WS (R^2^ difference = 0.124) and 11.4 percentage points for PS (ΔR^2^ = 0.114), indicating that eHF thresholds capture speech-relevant cochlear damage that is only partly reflected in conventional audiometry. Together with the steeper age-related deterioration of eHF PTA compared to PTA₄ (**Fig. 3a,b**), this pattern supports the clinical added value of incorporating eHF measures into routine assessment. In contrast, OAE amplitude at 5.273 kHz exhibited a linear relationship for WS and cubic for PS (WS: R^2^ = 0.196, linear *p* < 0.0001; PS: R^2^ = 0.219, cubic *p* < 0.0001; **Figure S2, Table S7**), indicating a roughly proportional decrement in speech scores as OHC function decreases. These findings suggest that, in this cohort and listening condition (+4 dB SNR), eHF is sufficient to index the cochlear pathology that matters for speech, and OHC-specific measures offer limited incremental value for prediction beyond eHF PTA; consistently, none of the OAE terms was significant in the multivariable models (**Table S9**). The corresponding relationships between WS/PS and ABR measures are illustrated in **Fig. 4c-f,i-l**: Wave I amplitude (Eclipse only) exhibited a shallow linear association with WS and PS (n = 134 ears; WS: R^2^ = 0.045, linear *p* = 0.014; quadratic *p* = 0.839; PS: R^2^ = 0.045, linear *p* = 0.014; quadratic *p* = 0.541; **Fig. 4c,i**), with no improvement from higher-order terms, indicating that amplitude contributes minimally to explaining speech variance. Wave I latency showed a linear association for WS (WS: R^2^ = 0.115, linear *p* <0.0001; **Fig. 4d**) and quadratic improvement for PS (PS: R^2^ = 0.115, quadratic *p* = 0.024; **Fig. 4j**), showing modest but significant association with both outcomes. In contrast, Wave III latency required cubic terms for optimal fit (WS: R^2^ = 0.088, cubic *p* = 0.0017; PS: R² = 0.112, cubic *p* < 0.0001; **Fig. 4e,k**), indicating strong nonlinearity, while Wave V latency showed only weak linear relationships (WS: R^2^ = 0.065, quadratic *p* = 0.868; PS: R^2^ = 0.058, quadratic *p* = 0.986; **Fig. 4f,l**, **Table S7**). Together, these polynomial fits indicate that eHF PTA is a dominant, at least in part, predictor of speech outcomes in this cohort, accounting for 34–36% of univariate variance in speech scores (substantially more than conventional audiometry or any ABR measure), yet leaving the majority of variance (∼65%) unexplained by peripheral auditory measures alone. The strong cubic relationships for eHF PTA and Wave III latency justify the use of higher-order terms in subsequent multivariable models, in which eHF PTA, OAE and Wave III latency entered with cubic terms, PTA₄ with a quadratic term, and Wave I and V latency as linear terms.

**Figure 4:**
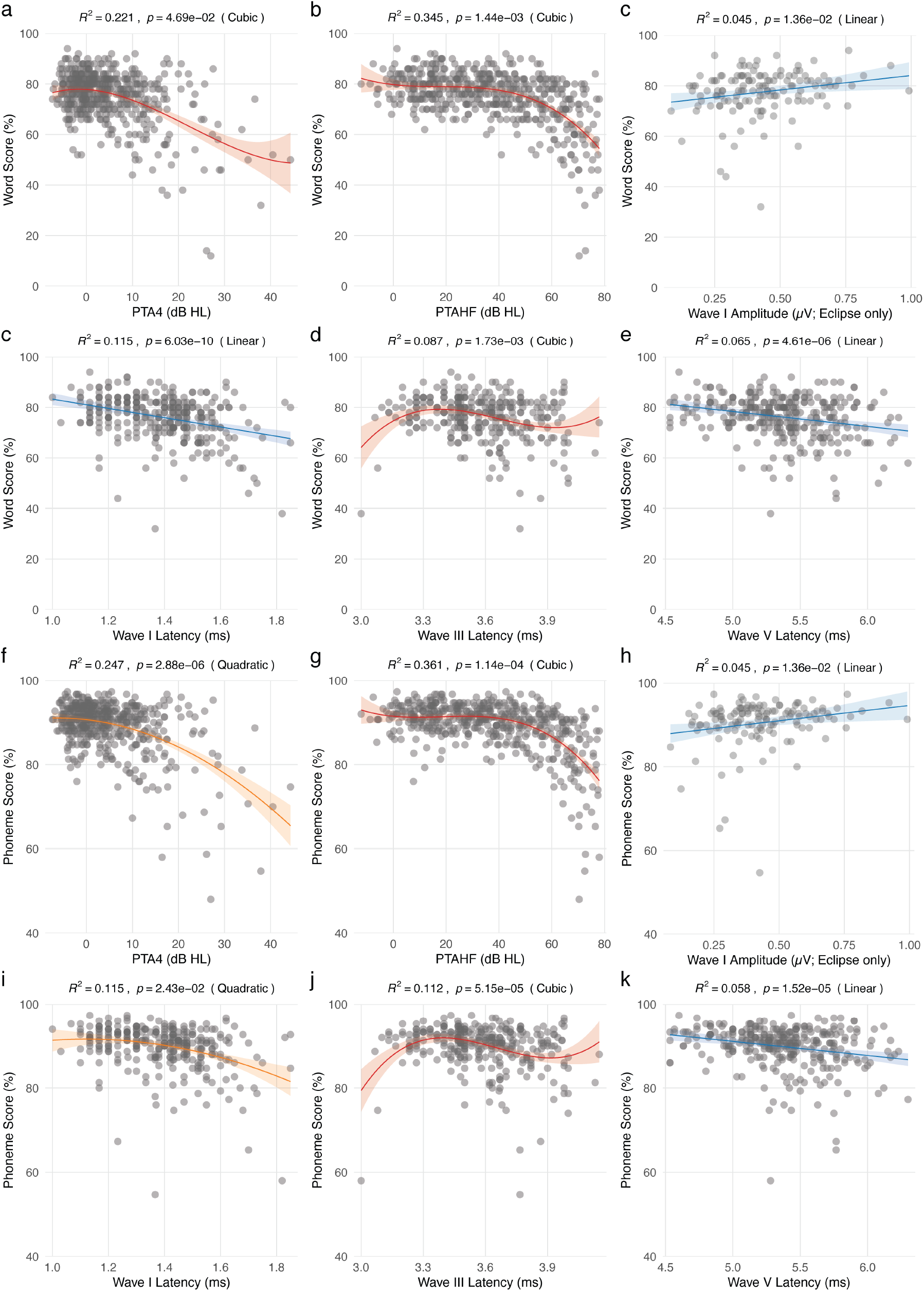
Univariate correlations with data-driven polynomial fits. Word score (WS) versus (a) PTA4, (b) EHF PTA, (c) Wave I amplitude (Eclipse only), and latencies of Wave I (d), III (e), and V (f). Phoneme score versus (g) PTA4, (h) EHF PTA, (i) Wave I amplitude (Eclipse only), and latencies of Wave I (j), III (k), and V (l). Best fits are reported for linear (blue), and quadratic (orange), and cubic (red) with shaded regions as 95% confidence intervals, and R^2^ and p values on top of the respective panels. Data relates to **Figure S2 and Table S7**.

### Incremental Contributions of Hearing Loss, ABR, and Interactions to Speech Models

Blockwise model progression analyses confirmed that each predictor domain made a significant incremental contribution to explaining variance in speech outcomes (**Table S8**). For WS, adding hearing loss measures to an age/gender baseline model increased R^2^ from 0.23 to 0.29 (DR^2^ = 0.064, *p* = 0.0016), with further gains when ABR latencies (R^2^ = 0.34, DR^2^ = 0.049, *p* = 0.0011) and the Age x Wave I latency interaction (R^2^ = 0.36, DR^2^ = 0.022, *p* = 0.0021) were included. PS showed a similar pattern but with larger improvements from hearing loss and ABR blocks: R² rose from 0.24 (age/gender only) to 0.33 with hearing loss (ΔR^2^ = 0.092, *p* = 9.7×10⁻⁶), to 0.40 with ABR latencies (DR^2^ = 0.067, *p* = 1.5×10⁻⁵), and to 0.42 after adding the interaction (DR^2^ = 0.016, *p* = 0.006). These stepwise gains indicate that peripheral hearing loss, brainstem timing, and age-dependent interactions each explain unique components of speech variance beyond demographics alone. In the Eclipse-only sequence (n = 122 ears), adding Wave I amplitude (WS: DR^2^ = 0.002, *p* = 0.57; PS: DR^2^ = 0.003, *p* = 0.44) and the Age x Wave I amplitude interaction (WS and PS: DR^2^ < 0.001, *p* ≥ 0.69) did not improve model fit (**Table S8**).

### Comparison of the mechanisms underlying WS and PS

Despite the fact that WS and PS are highly correlated (r = 0.934, *p* < 0.001), research suggest these two measures dissociate under specific hearing loss profiles [46], prompting to investigate further the differences between the two. Indeed, the apparent U-shaped age effect in univariate analysis in **Fig. 4** may mask underlying mechanisms. Once controlling for hearing loss and neural timing measures, WS shows genuine age-acceleration (b = −0.0101, 95% CI −0.018 to −0.002, *p* = 0.012; **Fig. 5a, Table S9**), indicating that age-related deterioration accelerates in older age. In contrast, PS showed only linear age effects without significant quadratic acceleration (age^2^ b = −0.0041, 95% CI −0.009 to 0.001, *p* = 0.113; **Table S9**). Specifically, with other predictors held at their mean, the model-implied WS decline was approximately 0.1 percentage points per year at age 50 and 0.6 percentage points per year at age 75, whereas the corresponding PS declines were smaller (approximately 0.04 and 0.25 points per year) and the quadratic term was not significant. This differential trajectory indicates that complex linguistic processing (word recognition) is more age-vulnerable than basic acoustic perception (phoneme identification).

**Figure 5.**
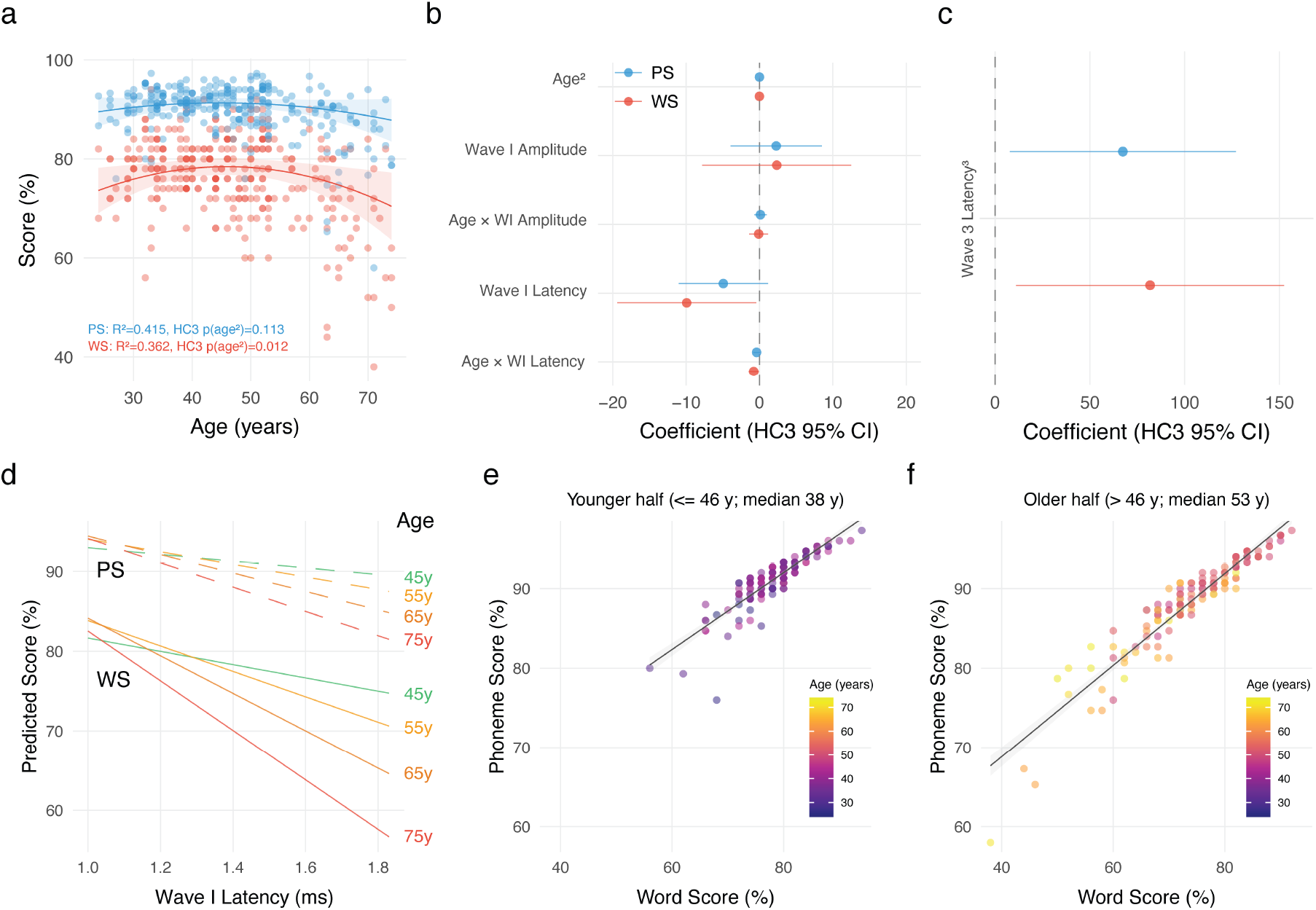
Comparison of the mechanisms underlying WS and PS. (a) WS (red) exhibits accelerating quadratic decline with age, while PS (blue) shows linear decline with age. Solid lines represent predicted values from multivariable models controlling for hearing loss, neural timing measures, outer hair cell function, and gender. Shaded regions indicate 95% confidence intervals. Open circles denote raw data points (n = 298 ears from 163 participants). (b-c) Forest plots comparing effect sizes (HC3 robust SE) between WS and PS for key predictors; Wave I amplitude and Age x Wave I amplitude are from the Eclipse-only model (n = 122 ears), all other terms from the latency model (n = 298 ears). Summary statistics are in **Tables S9 and S10**. (d) Four age groups (45, 55, 65, 75 years) show divergent trajectories as Wave I Latency (neural timing) increases (**Table S12**). Older adults (red, 75y) experience steeper Word Score decline (solid line) compared to Phoneme Score decline (dashed line). Young adults (green, 45y) show minimal decline across both measures. (e-f) High correlation (r = 0.934) masks differential aging patterns. Points colored by age (≤46 y in (e) and >46 y in (f), split at the median age of the complete-case subset) reveal that older adults (red) have relatively preserved PS but reduced WS, indicating dissociable age-related mechanisms (**Table S13**). Panels a–d control for age, age^2^, gender, low-frequency hearing loss (PTA4, PTA_4_^2^), high-frequency hearing loss (eHF PTA, eHF PTA^2^, eHF PTA^3^), outer hair cell function (OAE, OAE^2^, OAE^3^), Wave I Latency, Age x Wave I latency, Wave III Latency, Wave III Latency^2^, Wave III Latency^3^, and Wave V Latency. HC3 heteroscedasticity-consistent standard errors used to account for unequal variances in speech perception measures. Except for the Wave I amplitude terms, the sample was restricted to ears with complete data on age, WS, PS, PTA, OAE and ABR latency measures (complete-case subset, n = 298).

Multivariable polynomial regression models identified distinct predictor patterns for word versus phoneme recognition. Unstandardized coefficients showed that, contrary to predictions from the cochlear synaptopathy hypothesis [16–18] whereby lower Wave I amplitude is associated with speech recognition deficits [15, 20], Wave I amplitude, the putative neural marker of cochlear inner hair cell-auditory nerve synapse loss, showed no significant relationship to either WS (b = 2.35, 95% CI −7.81 to 12.52, *p* = 0.647) or PS (b = 2.28, 95% CI −3.95 to 8.51, *p* = 0.470; **Fig. 5b, Table S10**) in the Eclipse-only model (n = 122 ears from 68 participants), where Wave I amplitude reliability is excellent (ICC = 0.96) [28]. Furthermore, the Age x Wave I amplitude interaction was not significant for either outcome (WS: b = −0.110, 95% CI −1.386 to 1.165, *p* = 0.864; PS: b = 0.134, 95% CI −0.705 to 0.972, *p* = 0.752; **Fig. 5b**, **Table 2**). Collectively, these results indicate that ABR Wave I amplitude shows no detectable association with word or phoneme recognition in presence of background noise, even in the highest-quality, most reliable recording subset. In striking contrast, Wave I Latency that reflects more auditory nerve conduction velocity rather than neural synchrony demonstrated a predictive value for WS but not PS (WS: b = −9.92, 95% CI −19.40 to −0.45, *p* = 0.040; PS: b = −4.93, 95% CI −11.01 to 1.15, *p* = 0.112; **Fig. 5b, Table S9**). Each additional millisecond of Wave I latency was associated with approximately 10-point decline in Word Score and 5-point decline in Phoneme Score, with WS showing approximately twice the sensitivity to neural timing delays. Critically, the Age x Wave I latency interaction was substantially larger in magnitude for WS compared to PS (**Fig. 5b**). Specifically, WS showed a significant Age x Wave I latency interaction (b = −0.788, 95% CI −1.484 to −0.093, *p* = 0.027), indicating that older adults experience greater speech perception decline as Wave I latency increases (**Fig. 5b**, **Table 2, Table S9**). The PS interaction was not significant being substantially weaker in magnitude (b = −0.401, 95% CI −0.844 to 0.042, *p* = 0.076), with the WS age-interaction effect approximately 1.97 times the magnitude of the PS effect (coefficient ratio; **Fig. 5b**, **Table 2, Table S9**); the difference between outcomes was not formally tested. With participant-clustered (CR3) standard errors, the Age x Wave I latency interaction remained significant for WS (95% CI −1.51 to −0.06, *p* = 0.035) and non-significant for PS (*p* = 0.096), and Wave I amplitude remained unrelated to either outcome in the Eclipse-only model (WS: *p* = 0.65; PS: *p* = 0.50; **Table S11**). Adjustment for recording device did not alter the interaction (WS: b = −0.79, *p* = 0.027), although the Wave I latency main effect for WS became marginal (*p* = 0.061; **Table S11**). In the Eclipse-only model, the Age x Wave I latency estimate for WS was of similar magnitude (b = −0.83) but imprecise (95% CI −2.99 to 1.34, *p* = 0.450; **Table S10**). This dissociation suggests a differential age-dependent vulnerability: while both word and phoneme recognition are sensitive to peripheral neural transmission delays, the age-dependent amplification of neural timing vulnerability is approximately twice as pronounced for word recognition, suggesting task-specific mechanisms of aging. These findings collectively suggest that delayed neural transmission (reflected in increased Wave I latency), rather than synapse loss (reflected in decreased amplitude), is more closely associated with age-related speech perception decline, although the cross-sectional design precludes causal inference. The selective age-amplification of the Wave I latency effect for word recognition (vs. phoneme recognition) indicates that complex linguistic processing may depend more heavily on the precision of early neural timing information than basic acoustic-phonetic processing.

**Table 2.** Comparison of Age interactions with Wave I amplitude or latency on speech predictions. Significant *p* values are shown in bold.

| Interaction | Outcome | β | SE | 95% CI | p-value |
|---|---|---|---|---|---|
| Age x Wave I amplitude | WS | -0.11 | 0.64 | -1.39 to 1.17 | 0.864 |
| Age x Wave I amplitude | PS | 0.13 | 0.42 | -0.70 to 0.97 | 0.752 |
| Age x Wave I latency | WS | -0.79 | 0.35 | -1.48 to -0.09 | **0.027** |
| Age x Wave I latency | PS | -0.40 | 0.23 | -0.84 to 0.04 | 0.076 |

Age x Wave I amplitude estimates are from the Eclipse-only model (n = 122 ears; **Table S10**); Age x Wave I latency estimates are from the combined-device latency model (n = 298 ears; **Table S9**). β, unstandardized coefficient; SE, HC3 heteroscedasticity-robust standard error; CI, confidence interval.

Surprisingly, Wave III latency cubed, representing higher-order (cubic) nonlinearity in central auditory brainstem processing, significantly predicted both outcomes with large, positive effects (**Fig. 5c, Table S9**). Specifically, WS was predicted by Wave III Latency^3^ (b = 81.78, 95% CI 11.07 to 152.50, *p* = 0.024), and PS was similarly predicted (b = 67.42, 95% CI 7.74 to 127.11, *p* = 0.027; **Fig. 5c, Table S9**; participant-clustered CR3: WS, *p* = 0.049; PS, *p* = 0.051; **Table S11**). With participant clustering, inference for this term rested on few effectively informative participants (Satterthwaite df ∼ 11), so the cubic Wave III effect should be interpreted with caution. The comparable magnitude of Wave III Latency^3^ effects between outcomes (difference in coefficients = 14.36 points, approximately 18% relative difference) contrasts sharply with the substantially differential Age x Wave I latency effects (coefficient ratio = 1.97-fold difference). This dissociation explains that higher-order (cubic) central auditory processing effects impact word and phoneme recognition similarly, indicating shared reliance on central auditory complexity, whereas, in contrast, the age-dependent neural timing vulnerability, specifically the age-amplified impact of peripheral wave initiation delays (Age x Wave I latency), shows outcome-specific vulnerability that is unique to word-level processing. This pattern suggests that aging selectively compromises mechanisms dependent on early neural precision (Wave I timing), whereas cubic nonlinearities in central processing (Wave III cubic term) are inherently task-invariant. Finally, Wave V latency showed no significant associations in the multivariable model (WS: b = −0.673, 95% CI −3.591 to 2.246, *p* = 0.650; PS: b = −0.335, 95% CI −1.831 to 1.161, *p* = 0.660), indicating that the most distal brainstem response does not provide independent predictive value when upstream measures (Wave I latency, Wave III latency) are included in the model (**Table S9**).

### Age-Stratified Trajectories Reveal Selective Word Score Vulnerability

Predicted trajectories across four age strata (45, 55, 65, 75 years) reveal that while young adults (45y) maintain stable performance across Wave I latency values (WS predicted 74.7–81.7%, PS predicted 89.5–93.0%), older adults (75y) show marked declines as Wave I latency increases, with WS dropping to 56% and PS to 80% at the maximum observed Wave I latency (**Fig. 5d, Table S12**). This 27-percentage-point drop in WS (from 82.3% to 55.6%) versus 13-percentage-point drop in PS (from 93.8% to 80.4%) demonstrates that peripheral neural timing delay effects increase with advancing age, with word-level processing showing approximately 2.0 times greater vulnerability compared to phoneme recognition. The age-stratified trajectories visually demonstrate the Age x Wave I latency interaction. While young adults’ performance remains relatively insensitive to Wave I latency across the observed range, older adults show substantially steeper slopes, indicating that age unmasks or amplifies the neural timing dependency of speech recognition.

### Dissociation Between Word and Phoneme Recognition Across Age Groups

I tested whether word and phoneme scores show differential relationships with age and neural timing by constructing multivariable polynomial regression models with interaction terms. Despite high overall correlation (Pearson r = 0.934, *p* < 0.001), WS and PS show dissociable aging patterns and neural correlates with WS, but not PS, exhibiting accelerated age-related decline and age-amplified vulnerability to neural timing delays. Yet, both outcomes were equally driven by central processing measures as indicated by Wave III latencies (**Fig. 5c)**. This dual-process profile suggests that word recognition is preferentially vulnerable to age-related neurobiological changes and early neural timing deficits, while phoneme recognition remains more robust to age-dependent mechanisms, despite both being sensitive to central auditory complexity. Age-stratified analysis revealed selective word score vulnerability despite the high bivariate correlation. Young adults (≤ 46 years, the median age of the complete-case subset; n = 153 ears) clustered tightly in the upper-right quadrant (high WS, high PS) with a regression slope (0.48, 95% CI 0.42–0.54, *p* < 0.001; **Fig. 5e,f**). Older adults (older > 46 years, n=145 ears) showed greater scatter and were concentrated in the lower-left region (lower WS, lower PS) with a steeper slope (0.58, 95% CI 0.50–0.65, p < 0.001; **Fig. 5e,f**), although the difference in slopes did not reach significance (D = 0.09, 95% CI −0.001 to 0.19, *p* = 0.051). The steeper point estimate in older adults (0.58 vs 0.48) suggests stronger WS-PS coupling with age, whereby phoneme recognition changes more steeply relative to word score, a pattern compatible with differential age-related vulnerability of complex linguistic versus acoustic-phonetic processing.

## Discussion

This cross-sectional analysis of 526 ears from 263 tinnitus-free adults reveals three central findings regarding age-related speech perception decline. First, eHF PTA (10–16 kHz) is the dominant predictor of speech recognition, explaining 34–36% of variance in both word and phoneme scores, substantially more than conventional audiometry or any physiological measure. Second, ABR Wave I amplitude contributes no additional explanatory power, inconsistent with the “hidden hearing loss” hypothesis even when tested in speech-weighted noise (+4 dB SNR) where these effects should be most evident [13, 20, 47]. Third, Wave I latency reveals age-dependent vulnerability specific to complex linguistic tasks (word recognition), suggesting that neural conduction slowing, rather than synapse loss, may contribute to age-related speech deficits, though this finding requires validation. Collectively, these results support eHF audiometry as a primary clinical tool for predicting speech outcomes in aging populations and challenge the translational relevance of ABR-based synaptopathy indices.

Past studies have attempted to address the peripheral contributors to SPiN deficits, either focusing on eHF PTA [11], DPOAE [24, 25], or ABR markers [21] in isolation, or in small [26, 27], highly selected samples [20]. Of those rare studies that used eHF PTA, DPOAE and ABR in relation to SPiN [26, 27], these were not covering older age groups, and insufficient in sample size to detect the non-linearities presented herein. The present contribution is thus methodological (testing all mechanisms together) and population-based (all age groups with substantial audiometric variability). In the context of prior literature, our findings partially confirm those from Motlagh Zadeh et al. where eHF hearing enhances SPiN [11]. The linear worsening of eHF hearing with aging observed here is outstanding for a single auditory measure explaining 64% of the variance. Unlike other measures that show accelerating (quadratic) decline, eHF PTA declines linearly, steadily and predictably throughout the lifespan making it a reliable biomarker of cochlear aging far better than any other conventional audiometry, all below 30%. Since eHF PTA tracks cumulative cochlear damage with such fidelity, and since speech relies on intact cochlear function across the whole frequency spectrum, eHF PTA naturally emerges as the dominant speech predictor. Based on the differential age-related trajectories (eHF PTA R^2^ = 0.641 vs. PTA R^2^ = 0.158), mean eHF thresholds reach 25 dB HL (mild hearing loss) at around 40 years of age, whereas mean PTA₄ remains below this level across the entire age range sampled (20–84 years), providing a critical window for early intervention and hearing conservation counseling.

Our finding that eHF PTA (10–16 kHz) explains 34–36% of speech variance in univariate polynomial models (R^2^ = 0.34–0.36), substantially more than conventional PTA₄ (22–25%), aligns with emerging evidence that standard audiometry systematically underestimates cochlear damage in aging populations [11]. Speech contains substantial high-frequency acoustic cues extending into the 8–16 kHz range, with eHF energy accounting for approximately 10–20 dB below mid-frequency peaks when analyzed using physiologically-relevant equivalent rectangular bandwidth (ERB) filters [48, 49]. Fricative contrasts (/s/ vs. /ʃ/), stop consonant bursts, and voice pitch harmonics all carry linguistically important information in this range, with the maximum audible low-pass cutoff for speech reaching approximately 13 kHz in young normal-hearing listeners [48]. Critically, speech radiation patterns are frequency-dependent: eHF energy radiates primarily toward the front of a talker, whereas low-frequency energy is omnidirectional [50]. In ecological “cocktail party” scenarios where target talkers face the listener but interfering talkers are rotated away, access to eHFs (up to 20 kHz) significantly improves SRTs by 1.4–2.0 dB relative to speech low-pass filtered at 8 kHz [51]. This frequency-dependent directivity makes eHF energy a salient cue for auditory object formation, facilitating both detection and segregation of target talkers from background talkers [50, 51]. Traditional laboratory studies with all talkers facing the listener artificially equalize eHF energy across speech sources, obscuring these contributions [49, 52]. Our findings demonstrate that eHF loss predicts speech outcomes even in controlled (4 dB SNR) conditions, suggesting that cumulative loss of eHF phonetic information and reduced segregation cues is clinically meaningful across diverse acoustic environments. The high prevalence of undetected eHF loss among young adults with self-reported listening difficulties [53] supports extending routine audiometry beyond 8 kHz, particularly for individuals presenting with unexplained speech-in-noise complaints, ototoxicity exposure, or noise-related occupations.

The fact that ABR wave I amplitude did not account for variability in speech-in-noise performance contrasts with predictions that cochlear synaptopathy (CS) would be a primary driver of SPiN deficits [18, 21]. While animal studies convincingly demonstrate that noise exposure causes permanent loss of auditory nerve fiber (ANF) synapses with inner hair cells, leading to reduced ABR wave I without threshold elevation [16, 54, 18], human translation has proven challenging [22]. Recent human temporal bone studies provide direct anatomical evidence for age-related neural degeneration contributing to poor word-discrimination scores among individuals with similar audiometric thresholds [19, 55, 56]. However, multiple large-scale human studies have failed to find consistent relationships between putative physiological markers of CS and speech-in-noise performance [57–61]. The impact of CS on hearing may be most pronounced early in the pathological cascade, before substantial hair cell loss. Early-life noise exposure accelerates age-related cochlear synaptopathy in mice, with effects spreading from high-frequency regions initially affected to lower frequencies as animals age [62]. This noise-accelerated synaptopathy precedes threshold elevation by months, but once animals reach an advanced age (∼20 months post-exposure), threshold shifts and OHC loss emerge, converging with aging-only pathology. This suggests CS may be an early vulnerability marker, but its relative contribution to hearing impairment diminishes once threshold-elevating pathologies (e.g., OHC loss, strial degeneration) accumulate. Despite mixed behavioral evidence [57–61], computational models strongly support CS impairing supra-threshold temporal coding [13, 15, 47]. Gerbils with noise-induced CS display impaired neural discrimination of vowel-consonant-vowels in background noise at high sound levels (75 dB SPL), which was surprisingly improved at moderate levels (60 dB SPL), something reproduced in a phenomenological model [63]. A fundamental challenge is the inability to directly measure CS in living humans since all proposed markers are indirect and influenced by multiple cochlear pathologies. While CS undoubtedly occurs in aging and noise-exposed human cochleas, our study joins growing evidence that CS alone does not account for large inter-individual variability in speech-in-noise performance among individuals with clinically normal audiograms. Multiple peripheral pathologies (synaptopathy, demyelination, strial degeneration, OHC dysfunction) and central factors (cognitive control, attention, neural gain) likely interact to determine real-world listening abilities.

In contrast to Wave I amplitude, Wave I latency appears as a stronger predictor of speech pathology. The selective interaction between Wave I latency and age for word recognition (WS, *p* = 0.027) but not phoneme recognition (PS, *p* = 0.076) reflects differential temporal integration demands: word-level comprehension requires extended temporal windows (400-600 ms) encompassing multiple phonemes, making it vulnerable to conduction delays that accumulate across longer sequences, whereas phoneme recognition operates within brief acoustic windows (50-100 ms) where conduction delays have minimal impact. Indeed, this dissociation is consistent with hierarchical processing models wherein word-level comprehension requires precise temporal integration across multiple phonemes and syllables—a process that becomes increasingly vulnerable when peripheral timing delays compound with age-related central processing decline. In contrast, phoneme recognition relies on shorter temporal integration windows and benefits from high acoustic-phonetic redundancy, rendering it relatively robust to peripheral timing variability even in older age [64–67]. These findings suggest that peripheral neural timing precision, rather than synaptic loss (as measured by Wave I amplitude), represents the rate-limiting step for complex speech understanding in aging. These findings suggest that age-related speech deficits are more closely associated with slowed auditory nerve conduction than with cochlear synaptic loss inferred from Wave I amplitude. In support of these findings, acute demyelination of auditory nerve fibers in mice using a transient ablation of Schwann cells leads to a hidden hearing loss phenotype with normal thresholds with both a decrease in Wave 1 amplitude and increased Wave 1 latency in absence of synapse loss [68]. Our data in aging humans, however, reveal a different profile: ABR Wave I amplitude shows no association with speech-in-noise performance, whereas Wave I latency and its interaction with age were associated with word recognition, particularly in older adults, but not with phoneme recognition. This suggests that in heterogeneous human cohorts, the temporal imprecision resulting from decreased peripheral conduction may be more tightly coupled to complex speech deficits than the overall reduction in neural output, and that amplitude-based ‘synaptopathy’ markers may lack specificity for the age-related mechanisms at play.

WS appears to represent higher-level, plasticity-dependent speech comprehension, while PS reflects more fundamental, robust acoustic processing. This dissociation has important implications for age-based intervention strategies. Our best models explain 32-38% of speech variance, leaving ∼65% unexplained. What could account for this residual? Working memory, attention, processing speed, and executive function all contribute to speech understanding, especially in challenging conditions [69, 70]. Age-related cognitive decline may co-occur with hearing loss but follow an independent trajectory [71], and cognitive reserve may compensate for peripheral deficits [72]. Our models did not include cognitive assessments and thus future work incorporating measures like the Montreal Cognitive Assessment (MoCA) adapted to individuals with hearing impairment [73] or digit span could partition variance between sensory and cognitive domains. Cortical auditory processing, phonemic decoding, lexical access, predictive coding, is entirely unobserved in click ABRs, which reflects only the first 10 ms of the auditory pathway. Cortical evoked potentials (e.g., P1-N1-P2 complex, mismatch negativity) or speech-evoked envelope following responses may capture variance orthogonal to audiometry [58, 74]. Aging affects cortical temporal processing, neural noise, and speech-specific encoding [75], potentially explaining inter-individual differences in speech perception despite similar audiograms. In addition, vocabulary size, phonemic familiarity, native language experience, and lifetime auditory enrichment all modulate speech recognition [76, 77]. Comprehensive models integrating eHF audiometry, cognitive assessments, cortical evoked potentials, and speech-in-noise testing across diverse listening environments (reverberation, competing talkers, audiovisual) are needed to fully map the determinants of real-world communication ability in aging.

Women obtained modestly higher word and phoneme scores than men after adjustment for age, but gender did not contribute independently in the fully adjusted models, and there was no clear evidence that age-related trajectories of speech perception differed between women and men. These null interactions do not establish equivalence: women were over-represented (67.7%), which reduces precision for men and for interaction terms, and only self-reported gender was available, so sex-linked biological contributions to cochlear aging cannot be separated from gender-related exposures such as occupational noise.

## Strengths and Limitations

The main strengths of this study are its population-based design with a large, broadly-recruited cohort focusing on healthy tinnitus-free individuals (N = 263 adults across the adult lifespan), the use of comprehensive and multi-domain audiometric characterization (audiometry 0.125–16 kHz, OAE, click-ABR, and speech outcomes), rigorous quality control of ABR recordings (double-blind waveform identification, test-retest analyses), and the use of robust statistical models with pre-specified hierarchical structure and heteroscedasticity-consistent standard errors. However, the use of a tinnitus-focused cohort to study tinnitus-free individuals may introduce volunteer bias, since participants may have self-selected based on hearing concerns even in the absence of tinnitus. Other limitations include the cross-sectional design, which precludes causal inference about whether eHF loss precedes speech decline, something that would require larger efforts in population screens to answer this question. The restriction to Swedish-speaking, highly educated urban adults, limits generalization. Generalizability is further limited to tinnitus-free adults able to complete the full test battery, to the age range sampled (20–84 years), and to the specific transducers, stimulus protocols and device output limits used here; the over-representation of women (67.7%) also limits precision for men. Analyses were performed at the ear level, although participant-clustered standard errors did not change the main conclusions (the Wave III latency³ effect on PS became borderline, p = 0.051), the two ears of a participant are not independent observations. The complete-case multivariable sample (298 of 526 ears) may differ from the full cohort, and Wave I amplitude could only be analyzed in the smaller Eclipse-only subset (122 ears), which limits precision for amplitude and its interaction with age. In addition, the use of a single, moderately challenging SNR (+4 dB) may not reveal synaptopathy effects present at lower SNRs [13]. Finally, the high WS-PS correlation (r = 0.934) means that most main effects are inevitably replicated across both outcomes, however, the incremental scientific value justifying the retention of both measures resides primarily in the differential Age and Wave I Latency interaction.

## Implementation of Extended Audiometry in Clinical Practice and Clinical Trials

Whether in clinical practice or in clinical trials, implementation of the eHF measures may differ depending on the purpose. In clinical practice, standard pure-tone audiometry covers 0.125–8 kHz and is performed with commercially available, CE-marked audiometers. Extending the range to 10, 12.5, 14 and 16 kHz requires an audiometer with extended high-frequency capability together with a transducer calibrated for that range according to ISO 389-5 (for instance the Sennheiser HDA 200, which is widely available), a configuration offered by many current clinical audiometers, including the Madsen Astera^2^ used here. The marginal cost is therefore the examiner’s time, approximately 5–10 additional minutes per ear, rather than new hardware, which makes routine eHF screening feasible in clinical audiology, occupational health and hearing conservation programs. Since the purpose in this setting is to identify individuals who may have speech-in-noise difficulties rather than to quantify a threshold exactly, a device-limited “no response at maximum output” remains clinically informative, and the reduced maximum output at 14 and 16 kHz apparent in the older age groups (> 55 years; **Fig. 2a**) does not compromise the screening decision. However, age-related reference values remain incomplete above 8 kHz, since ISO 7029:2017 gives expected medians only up to 12.5 kHz and for information only, so interpretation at 14 and 16 kHz has to rely on published normative data (Motlagh Zadeh et al. [11], supplementary materials). In resource-limited settings, eHF screening could be prioritized for individuals with unexplained speech-in-noise complaints, a history of ototoxic exposure, or occupational noise exposure, where the clinical yield is likely to be highest. For multi-site clinical trials, the requirements may differ because the measurement must be comparable across sites and over time rather than merely informative for one patient. The maximum output attainable at 14 and 16 kHz is substantially lower than at conventional frequencies and varies with the audiometer–transducer combination, so thresholds become censored at the device limit, not that of the ear. Whereas I acknowledge that identical equipment across centers is rarely achievable in investigator-initiated trials, this wouldn’t be critical within a single site where censoring is a constant, however, across sites equipped differently, the censoring would shift between centers, and pooled eHF means, prevalences and change scores would reflect more the distribution of equipments’ ceiling effect rather than that of hearing. Such details need to be evaluated during study design, prior the first enrollment.

In conclusion, the present study confirms eHF pure-tone audiometry as a dominant clinical predictor of speech recognition performance and challenges the hypothesis that cochlear synaptopathy, as reflected in ABR wave I amplitude changes, contributes meaningfully to speech deficits in this large population. In contrast, ABR wave I latency, a marker of peripheral neural conduction timing, shows an age-dependent association with word recognition, particularly in older adults, suggesting that slowed neural timing rather than reduced response amplitude is more tightly linked to complex speech decline in aging. Routine extension of hearing assessment beyond 8 kHz may provide clinically valuable information regarding speech prediction and communication prognosis, particularly in older adults and those with noise exposure history. Future studies examining hearing–cognition relationships should routinely include eHF audiometry, alongside latency-based brainstem timing measures, to avoid an incomplete characterization of hearing status.

## Acknowledgments

I am indebted to Jonathon Whitton and Golbarg Mehraei (Decibel Therapeutics Inc.), Hugo Westerlund and Constance Leineweber (SLOSH) and Nancy Pedersen (LG), Inger Uhlen and Andra Lazar (Karolinska Hospital) for their collaboration, support and dedication to STOP. I am very thankful to Otometrics (Clément Sanchez, Aud) and Interacoustics (Stefan Pettersson, Diatec Sweden AB) for sharing their equipment throughout the project duration. I warmly thank Niklas K. Edvall who collected the majority of the STOP data, and also Rickard Redgård, Anders Sjöstrand, and Laura Inan for their contributions in the acquisition of auditory data. I express my gratitude to all other collaborators of TINNET, ESIT, UNITI who guided in the initial steps of STOP, including Rilana Cima, Birgit Mazurek, and Jose Antonio Lopez-Escamez. Particular thanks to Jan Bulla for his input on statistical models. I would like to thank all the participants who joined the project and supported this study, as well as staff from the Department of Physiology and Pharmacology, Karolinska Institutet.

## Data availability

Anonymized individual participant data (audiometry, OAE, ABR, speech scores) will be soon made available upon approval by the Swedish ethics regulations.

## Code availability

Statistical analysis scripts (R) and figure generation code are available at https://github.com/translational-audiology-lab/STOP_Speech

## Author Contributions

C.R.C. designed and directed the research, analyzed the data, generated the figures and tables, and wrote the manuscript.

## Use of AI

A large language model [Perplexity, Version 2.260428.0 (670)] was used during the drafting phase to assist in proofreading and revising author-written text, and verification that all cited numbers in the main text were consistent with the ones provided in the Supplementary materials. No AI tools were used to generate data, analyze data, produce figures, or create artwork. All content was subsequently written, revised, and verified by the author. The author takes full responsibility for the integrity, accuracy, and originality of all manuscript content.

