## Supplementary material for "Peripheral Predictors of Speech in Noise Deficits in a Population Cohort"

### Supplementary Materials

Supplementary Figures 1–2 and Tables S1–S13

#### Supplementary Figure 1

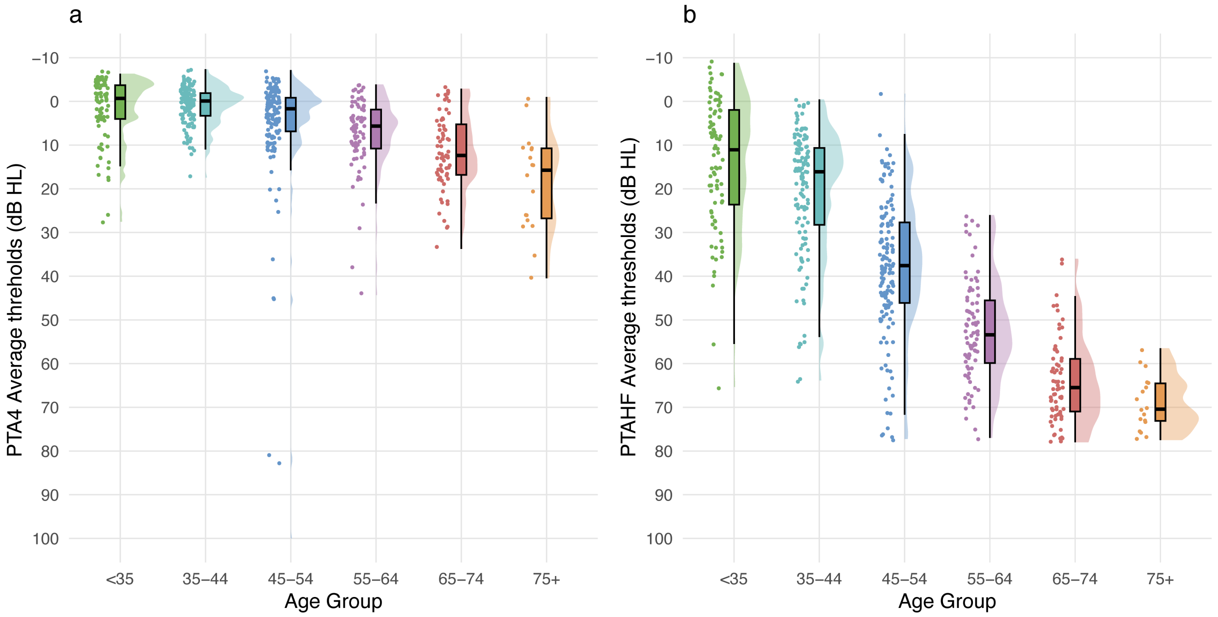


Age-stratification for PTA4 average (a) and eHF PTA (b). Summary statistics are shown as median and IQR (middle) and representation of distribution with one-sided violin plots (right).Age groups are displayed in different colors: >35 (green), 35-44 (light blue); 45-54 (blue); 55-64 (violet), 65-74 (red), and >75 (orange).

#### Supplementary Figure 2

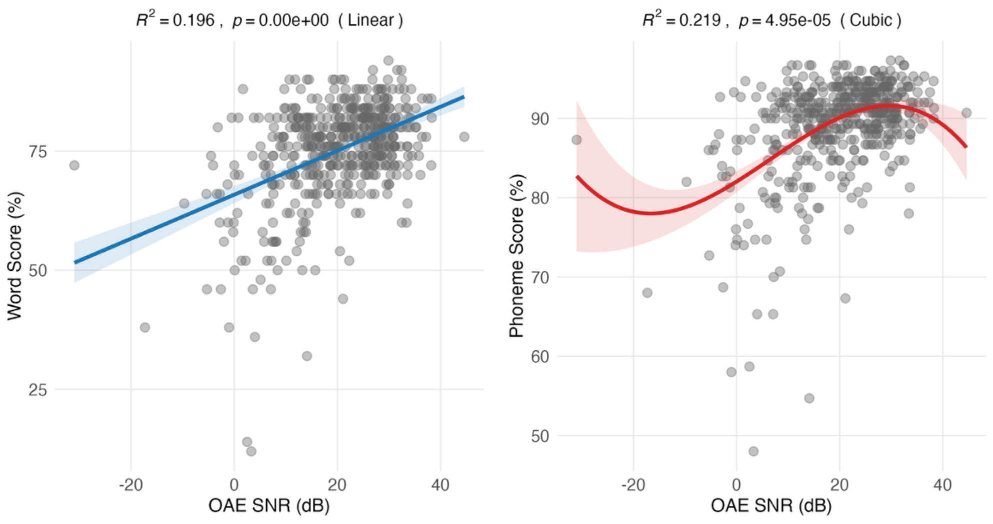


Word score (WS, left) and Phoneme Score (PS, right) versus OAE SNR. Best fits are reported for linear (blue) and cubic (red) with shaded regions as 95% confidence intervals, and R2 and p values on top of the respective panels.

### Supplementary Tables Outline


| Table | Title |
|---|---|
| Table S1 | Sociodemographics and disease prevalence in non-tinnitus participants from STOP |
| Table S2 | Audiometric values stratified by age groups |
| Table S3 | Audiometric values stratified by gender |
| Table S4 | Audiometric values stratified by eHF tertile |
| Table S5 | Age-related trajectories of audiometric, OAE, speech, and ABR measures (LRT-based polynomial models) |
| Table S6 | Age-by-gender interaction effects on speech perception and auditory measures |
| Table S7 | Univariate polynomial associations between hearing/auditory measures and speech scores (WS, PS) LRT-based model selection |
| Table S8 | Hierarchical model progression for combined-device latency and Eclipse-only amplitude models |
| Table S9 | Combined-device latency model coefficients with HC3-robust inference |
| Table S10 | Eclipse-only Wave I amplitude model coefficients with HC3-robust inference |
| Table S11 | Sensitivity analyses: HC3 versus participant-clustered (CR3) inference and adjustment for recording device |
| Table S12 | Combined-device latency model predictions with HC3-robust uncertainty |
| Table S13 | Age-stratified WS–PS correlations and HC3-robust regression slopes |

#### Table S1: Sociodemographics and disease prevalence in non-tinnitus participants from STOP

| Characteristics | No (%) |
|---|---|
| Total | 263 (100) |
| Gender | |
| Man | 85 (32.3) |
| Woman | 178 (67.7) |
| Education | |
| Middle School | 1 (0.4) |
| High School | 37 (14.1) |
| University | 197 (74.9) |
| Other | 28 (10.7) |
| Ear Diagnosis | Missing 32 |
| Acoustic Trauma | 1 (1.3) |
| Ear Barotrauma | 0 (0) |
| Presbycusis | 7 (3) |
| Sudden Hearing Loss | 2 (0.9) |
| Other Hearing Loss | 7 (3) |
| Menière's Disease | 0 (0) |
| Acoustic Neuroma | 0 (0) |
| Acute Otitis | 14 (6.1) |
| Serous Otitis | 4 (1.7) |
| Chronic Otitis | 3 (1.3) |
| Otosclerosis | 0 (0) |
| Other | 32 (13.9) |
| None | 172 (74.5) |
| Procedures | |
| Ear Surgery | 8 (3.5) |
| Dental Surgery | 69 (29.9) |
| Neurosurgery | 0 (0) |
| Lumbar puncture | 6 (2.6) |
| Chemotherapy | 5 (2.2) |
| Head and neck radiotherapy | 5 (2.2) |
| Electroconvulsive therapy | 0 (0) |
| Other | 22 (9.5) |
| None | 137 (59.1) |
| Devices | |
| Hearing aid | 0 (0) |
| Cochlear Implant | 0 (0) |
| Sound generator | 0 (0) |
| Combination | 0 (0) |
| None | 229 (99.1) |
| Pain Syndromes | |
| Headache | 28 (12.1) |
| Neck pain | 31 (13.4) |
| Ear pain | 3 (1.3) |
| TMJ | 6 (2.6) |
| Pain in the face | 0 (0) |
| Other | 11 (4.8) |

| Characteristics | No (%) |
|---|---|
| None | 171 (74) |
| Diagnosis | |
| TMJD | 0 (0) |
| Dental problems | 11 (4.8) |
| Meningitis | 2 (0.9) |
| MS | 0 (0) |
| Epilepsy | 1 (0.4) |
| Stroke | 1 (0.4) |
| Other cerebrovascular disease | 0 (0) |
| Dementia | 0 (0) |
| Other neurologic disease | 3 (1.3) |
| Anxiety | 15 (6.5) |
| Depression | 18 (7.8) |
| Emotional Trauma | 4 (1.7) |
| Excessive stress | 13 (5.6) |
| Difficulty faling asleep | 18 (7.8) |
| Difficulty staying asleep | 4 (1.7) |
| Low blood pressure | 13 (5.6) |
| High blood pressure | 16 (6.9) |
| Heart attack | 19 (8.2) |
| Thyroid disorder | 13 (5.6) |
| Diabetes | 20 (8.6) |
| Hyperinsulinemia | 2 (0.9) |
| Increased cholesterol | 17 (7.4) |
| Rheumatoid athritis | 4 (1.7) |
| Systemic lupus erythematosus | 0 (0) |
| Chronic synusitis | 1 (0.4) |
| Nasal septum deviation | 1 (0.4) |
| Syphilis | 0 (0) |
| HIV | 0 (0) |
| Lyme disease | 22 (9.5) |
| Anaemia | 11 (4.8) |
| Balance disorders | 5 (2.2) |
| Acid/gastroesophageal reflux | 11 (4.7) |
| Globus hystericus | 0 (0) |
| Other | 17 (7.4) |
| None | 118 (51.1) |

#### Table S2: Audiometric values stratified by age groups

| Variable | <35 | 35-44 | 45-54 | 55-64 | 65-74 | 75+ |
|---|---|---|---|---|---|---|
| N (ears) | 86 | 128 | 144 | 82 | 68 | 18 |
| Age (years) | 30.6 +/- 3.4 | 39.8 +/- 2.9 | 49.4 +/- 2.8 | 59.7 +/- 2.7 | 69.4 +/- 2.7 | 78.3 +/- 2.9 |
| Woman (%) | 56 (65%) | 86 (67%) | 100 (69%) | 62 (76%) | 40 (59%) | 12 (67%) |
| PTA4 (0.5-4 kHz) dB HL | 1.6 +/- 7.3 | 0.9 +/- 4.4 | 5.4 +/- 14.5 | 7.4 +/- 8.3 | 11.6 +/- 8.5 | 18.5 +/- 11.3 |
| eHF PTA (10-16 kHz) dB HL | 13.8 +/- 15.2 | 20.6 +/- 14.3 | 37.9 +/- 15.6 | 52.5 +/- 11.5 | 64.0 +/- 9.7 | 69.0 +/- 6.2 |
| LDL 8kHz (dB HL) | 75.6 +/- 16.4 | 79.6 +/- 18.1 | 86.4 +/- 17.5 | 88.4 +/- 12.9 | 93.9 +/- 9.6 | 90.6 +/- 12.7 |
| OAE at 5273 Hz (dB SNR) | 24.5 +/- 8.4 | 24.0 +/- 8.3 | 20.3 +/- 9.2 | 17.9 +/- 8.4 | 10.5 +/- 9.7 | 7.9 +/- 10.1 |
| Word score (%) | 78.3 +/- 6.5 | 79.0 +/- 5.8 | 77.8 +/- 7.3 | 71.9 +/- 10.4 | 66.9 +/- 11.0 | 55.6 +/- 19.4 |
| Phoneme score (%) | 90.9 +/- 3.7 | 91.5 +/- 2.9 | 90.9 +/- 3.7 | 87.1 +/- 7.0 | 84.8 +/- 6.7 | 77.8 +/- 12.3 |
| Wave I amplitude - Eclipse (uV) | 0.5 +/- 0.2 | 0.5 +/- 0.2 | 0.4 +/- 0.2 | 0.3 +/- 0.1 | 0.3 +/- 0.1 | - |
| Wave I latency (ms) | 1.3 +/- 0.1 | 1.3 +/- 0.2 | 1.4 +/- 0.1 | 1.5 +/- 0.1 | 1.5 +/- 0.2 | - |
| Wave III latency (ms) | 3.5 +/- 0.2 | 3.5 +/- 0.2 | 3.6 +/- 0.2 | 3.7 +/- 0.2 | 3.7 +/- 0.3 | - |
| Wave V latency (ms) | 5.3 +/- 0.3 | 5.3 +/- 0.3 | 5.4 +/- 0.4 | 5.5 +/- 0.4 | 5.6 +/- 0.3 | - |
| I-V inter-wave latency (ms) | 3.9 +/- 0.3 | 3.9 +/- 0.3 | 4.0 +/- 0.3 | 4.1 +/- 0.3 | 4.1 +/- 0.3 | - |

#### Table S3: Audiometric values stratified by gender

| Variable | Man | Woman |
|---|---|---|
| N (ears) | 170 | 356 |
| Age (years) | 49.6 +/- 14.0 | 49.0 +/- 13.5 |
| Woman (%) | 0 (0%) | 356 (100%) |
| PTA4 (0.5-4 kHz) dB HL | 6.2 +/- 10.9 | 4.8 +/- 10.6 |
| eHF PTA (10-16 kHz) dB HL | 38.3 +/- 23.3 | 35.6 +/- 22.1 |
| LDL 8kHz (dB HL) | 88.4 +/- 14.3 | 82.4 +/- 17.6 |
| OAE at 5273 Hz (dB SNR) | 16.7 +/- 10.3 | 21.3 +/- 9.6 |
| Word score (%) | 73.8 +/- 11.0 | 75.7 +/- 9.9 |
| Phoneme score (%) | 88.5 +/- 7.2 | 89.5 +/- 5.4 |
| Wave I amplitude - Eclipse (uV) | 0.4 +/- 0.2 | 0.4 +/- 0.2 |
| Wave I latency (ms) | 1.4 +/- 0.2 | 1.4 +/- 0.2 |
| Wave III latency (ms) | 3.6 +/- 0.2 | 3.5 +/- 0.2 |
| Wave V latency (ms) | 5.5 +/- 0.4 | 5.3 +/- 0.4 |
| I-V inter-wave latency (ms) | 4.1 +/- 0.3 | 3.9 +/- 0.3 |

#### Table S4: Audiometric values stratified by eHF tertile

| Variable | Low eHF loss | Medium eHF loss | High eHF loss |
|---|---|---|---|
| N (ears) | 176 | 175 | 175 |
| Age (years) | 37.2 +/- 7.1 | 48.0 +/- 8.8 | 62.4 +/- 10.6 |
| Woman (%) | 126 (72%) | 117 (67%) | 113 (65%) |
| PTA4 (0.5-4 kHz) dB HL | 0.0 +/- 5.1 | 3.2 +/- 5.4 | 12.5 +/- 14.3 |
| eHF PTA (10-16 kHz) dB HL | 10.9 +/- 7.8 | 36.0 +/- 6.9 | 62.6 +/- 8.5 |
| LDL 8kHz (dB HL) | 77.3 +/- 17.5 | 85.9 +/- 16.4 | 90.0 +/- 13.8 |
| OAE at 5273 Hz (dB SNR) | 25.8 +/- 7.3 | 21.3 +/- 8.0 | 12.4 +/- 9.8 |
| Word score (%) | 79.3 +/- 6.2 | 77.5 +/- 6.5 | 68.2 +/- 13.0 |
| Phoneme score (%) | 91.4 +/- 3.4 | 90.8 +/- 3.2 | 85.2 +/- 8.1 |
| Wave I amplitude - Eclipse (uV) | 0.5 +/- 0.1 | 0.4 +/- 0.2 | 0.3 +/- 0.1 |
| Wave I latency (ms) | 1.3 +/- 0.2 | 1.4 +/- 0.1 | 1.5 +/- 0.2 |
| Wave III latency (ms) | 3.5 +/- 0.2 | 3.6 +/- 0.2 | 3.7 +/- 0.2 |
| Wave V latency (ms) | 5.3 +/- 0.3 | 5.3 +/- 0.3 | 5.6 +/- 0.4 |
| I-V inter-wave latency (ms) | 3.9 +/- 0.3 | 4.0 +/- 0.3 | 4.1 +/- 0.3 |

### Table S5: Age-related trajectories of audiometric, OAE, speech, and ABR measures (LRT-based polynomial models)

| Variable | n | R2_linear | p_linear | R2_quadratic | p_quadratic | R2_cubic | p_cubic | Variable | Best_fit_model | Best_R2 | Decision_rule |
|---|---|---|---|---|---|---|---|---|---|---|---|
| PTA4 | 526 | 0.1451 | < 1e-16 | 0.1575 | 5.65e-03 | 0.1576 | 7.87e-01 | PTA4 | Quadratic | 0.1575 | Quadratic best (p_quad=0.0057 < 0.05, p_cubic=0.7875 ,> 0.05) |
| eHF PTA | 526 | 0.6407 | < 1e-16 | 0.6409 | 5.48e-01 | 0.6456 | 9.16e-03 | PTAHF | Linear | 0.6407 | Linear (p_quad=0.5481 ,> 0.05) |
| OAE | 526 | 0.2272 | < 1e-16 | 0.2372 | 9.00e-03 | 0.2373 | 7.89e-01 | OAE | Quadratic | 0.2372 | Quadratic best (p_quad=0.0090 < 0.05, p_cubic=0.7893 ,> 0.05) |
| WS | 511 | 0.2277 | < 1e-16 | 0.2965 | 6.00e-12 | 0.2965 | 9.53e-01 | WS | Quadratic | 0.2965 | Quadratic best (p_quad=0.0000 < 0.05, p_cubic=0.9529 ,> 0.05) |
| PS | 511 | 0.2140 | < 1e-16 | 0.2826 | 9.65e-12 | 0.2833 | 4.89e-01 | PS | Quadratic | 0.2826 | Quadratic best (p_quad=0.0000 < 0.05, p_cubic=0.4886 ,> 0.05) |
| Wave I amplitude - Eclipse | 140 | 0.1948 | 4.81e-08 | 0.1949 | 8.93e-01 | 0.2080 | 1.36e-01 | W1.Amplitude | Linear | 0.1948 | Linear (p_quad=0.8927 ,> 0.05) |
| Wave I latency | 324 | 0.1337 | 1.11e-11 | 0.1530 | 7.16e-03 | 0.1589 | 1.34e-01 | W1.Latency | Quadratic | 0.1530 | Quadratic best (p_quad=0.0072 < 0.05, p_cubic=0.1343 ,> 0.05) |
| Wave III latency | 322 | 0.0905 | 3.70e-08 | 0.1015 | 4.83e-02 | 0.1057 | 2.28e-01 | W3.Latency | Quadratic | 0.1015 | Quadratic best (p_quad=0.0483 < 0.05, p_cubic=0.2276 ,> 0.05) |
| Wave V latency | 321 | 0.1135 | 5.83e-10 | 0.1269 | 2.79e-02 | 0.1285 | 4.44e-01 | W5.Latency | Quadratic | 0.1269 | Quadratic best (p_quad=0.0279 < 0.05, p_cubic=0.4442 ,> 0.05) |

#### Table S6: Age-by-Gender interaction effects on speech perception and auditory measures

| model | outcome | term | estimate | std.error | statistic | p.value | conf.low | conf.high |
|---|---|---|---|---|---|---|---|---|
| 1.1_Age | PS | (Intercept) | 89.2076 | 0.2369 | 376.4943 | 0.00e+00 | 88.7421 | 89.6731 |
| 1.1_Age | PS | Age_c | -0.2036 | 0.0173 | -11.771 | 1.86e-28 | -0.2376 | -0.1696 |
| 1.4_Gender | PS | (Intercept) | 88.5006 | 0.4731 | 187.0509 | 0.00e+00 | 87.5711 | 89.4302 |
| 1.4_Gender | PS | GenderWoman | 1.0352 | 0.5725 | 1.8082 | 7.12e-02 | -0.0896 | 2.16 |
| 1.2_Age_quad | PS | (Intercept) | 90.6461 | 0.3064 | 295.8331 | 0.00e+00 | 90.0441 | 91.2481 |
| 1.2_Age_quad | PS | Age_c | -0.1703 | 0.0172 | -9.8911 | 3.25e-21 | -0.2041 | -0.1365 |
| 1.2_Age_quad | PS | Age_sq | -0.0077 | 0.0011 | -6.9737 | 9.65e-12 | -0.0098 | -0.0055 |
| 1.3_Age_cubic | PS | (Intercept) | 90.6707 | 0.3086 | 293.7986 | 0.00e+00 | 90.0644 | 91.277 |
| 1.3_Age_cubic | PS | Age_c | -0.1892 | 0.0323 | -5.8582 | 8.43e-09 | -0.2527 | -0.1258 |
| 1.3_Age_cubic | PS | Age_sq | -0.008 | 0.0012 | -6.6578 | 7.24e-11 | -0.0104 | -0.0056 |
| 1.3_Age_cubic | PS | Age_cu | 0 | 0.0001 | 0.693 | 4.89e-01 | -0.0001 | 0.0002 |
| 1.5_Age_Gender | PS | (Intercept) | 88.5264 | 0.4196 | 210.9534 | 0.00e+00 | 87.7019 | 89.3509 |
| 1.5_Age_Gender | PS | Age_c | -0.2034 | 0.0172 | -11.7914 | 1.56e-28 | -0.2373 | -0.1695 |
| 1.5_Age_Gender | PS | GenderWoman | 0.9974 | 0.5078 | 1.9643 | 5.00e-02 | -0.0002 | 1.9951 |
| 1.6_Age_x_Gender | PS | (Intercept) | 88.5321 | 0.4187 | 211.4534 | 0.00e+00 | 87.7095 | 89.3546 |
| 1.6_Age_x_Gender | PS | Age_c | -0.248 | 0.0298 | -8.3287 | 7.68e-16 | -0.3065 | -0.1895 |
| 1.6_Age_x_Gender | PS | GenderWoman | 0.9931 | 0.5066 | 1.9603 | 5.05e-02 | -0.0022 | 1.9884 |
| 1.6_Age_x_Gender | PS | Age_c:GenderWoman | 0.067 | 0.0365 | 1.8367 | 6.68e-02 | -0.0047 | 0.1387 |
| 1.1_Age | WS | (Intercept) | 75.0763 | 0.3991 | 188.1296 | 0.00e+00 | 74.2923 | 75.8603 |
| 1.1_Age | WS | Age_c | -0.3569 | 0.0291 | -12.2511 | 2.02e-30 | -0.4141 | -0.2996 |
| 1.4_Gender | WS | (Intercept) | 73.7901 | 0.8036 | 91.8292 | 6.10e-319 | 72.2114 | 75.3688 |
| 1.4_Gender | WS | GenderWoman | 1.8832 | 0.9723 | 1.9368 | 5.33e-02 | -0.0271 | 3.7935 |
| 1.2_Age_quad | WS | (Intercept) | 77.5221 | 0.5156 | 150.3555 | 0.00e+00 | 76.5092 | 78.5351 |
| 1.2_Age_quad | WS | Age_c | -0.3002 | 0.029 | -10.3644 | 5.80e-23 | -0.3571 | -0.2433 |
| 1.2_Age_quad | WS | Age_sq | -0.013 | 0.0018 | -7.0465 | 6.00e-12 | -0.0167 | -0.0094 |
| 1.3_Age_cubic | WS | (Intercept) | 77.5257 | 0.5195 | 149.2176 | 0.00e+00 | 76.5049 | 78.5464 |
| 1.3_Age_cubic | WS | Age_c | -0.3029 | 0.0544 | -5.5716 | 4.10e-08 | -0.4098 | -0.1961 |
| 1.3_Age_cubic | WS | Age_sq | -0.0131 | 0.002 | -6.4658 | 2.37e-10 | -0.0171 | -0.0091 |
| 1.3_Age_cubic | WS | Age_cu | 0 | 0.0001 | 0.059 | 9.53e-01 | -0.0002 | 0.0002 |
| 1.5_Age_Gender | WS | (Intercept) | 73.8353 | 0.7063 | 104.5336 | 0.00e+00 | 72.4476 | 75.223 |
| 1.5_Age_Gender | WS | Age_c | -0.3565 | 0.029 | -12.2797 | 1.56e-30 | -0.4135 | -0.2994 |
| 1.5_Age_Gender | WS | GenderWoman | 1.8171 | 0.8547 | 2.126 | 3.40e-02 | 0.1379 | 3.4962 |
| 1.6_Age_x_Gender | WS | (Intercept) | 73.8411 | 0.7062 | 104.561 | 0.00e+00 | 72.4536 | 75.2285 |
| 1.6_Age_x_Gender | WS | Age_c | -0.4017 | 0.0502 | -7.997 | 8.70e-15 | -0.5003 | -0.303 |
| 1.6_Age_x_Gender | WS | GenderWoman | 1.8127 | 0.8545 | 2.1213 | 3.44e-02 | 0.1338 | 3.4915 |
| 1.6_Age_x_Gender | WS | Age_c:GenderWoman | 0.0678 | 0.0615 | 1.1022 | 2.71e-01 | -0.0531 | 0.1887 |

#### Table S7: Univariate polynomial associations between hearing/auditory measures and speech scores (WS, PS) LRT-based model selection

| Outcome | Predictor | n | R2_linear | p_linear | R2_quadratic | p_quadratic | R2_cubic | p_cubic | Best_model | Best_R2 | Decision_rule |
|---|---|---|---|---|---|---|---|---|---|---|---|
| WS | PTA4 | 511 | 0.1961 | 0.00e+00 | 0.2152 | 4.65e-04 | 0.2213 | 4.69e-02 | Cubic | 0.2213 | Cubic (p_quad=0.0005 |
| WS | eHF PTA | 511 | 0.2531 | 0.00e+00 | 0.3316 | 6.27e-14 | 0.3448 | 1.44e-03 | Cubic | 0.3448 | Cubic (p_quad=0.0000 |
| WS | OAE at 5273 Hz | 511 | 0.1960 | 0.00e+00 | 0.2016 | 5.96e-02 | 0.2277 | 4.06e-05 | Linear | 0.1960 | Linear (p_quad=0.0596) |
| WS | Wave I amplitude - Eclipse | 134 | 0.0453 | 1.36e-02 | 0.0456 | 8.39e-01 | 0.0456 | 9.62e-01 | Linear | 0.0453 | Linear (p_quad=0.8387) |
| WS | Wave I latency | 317 | 0.1147 | 6.03e-10 | 0.1254 | 5.10e-02 | 0.1254 | 9.96e-01 | Linear | 0.1147 | Linear (p_quad=0.0510) |
| WS | Wave III latency | 315 | 0.0371 | 5.90e-04 | 0.0582 | 8.63e-03 | 0.0875 | 1.73e-03 | Cubic | 0.0875 | Cubic (p_quad=0.0086 |
| WS | Wave V latency | 314 | 0.0652 | 4.61e-06 | 0.0653 | 8.68e-01 | 0.0654 | 8.12e-01 | Linear | 0.0652 | Linear (p_quad=0.8676) |
| PS | PTA4 | 511 | 0.2139 | 0.00e+00 | 0.2471 | 2.88e-06 | 0.2499 | 1.68e-01 | Quadratic | 0.2471 | Quadratic (p_quad=0.0000 |
| PS | eHF PTA | 511 | 0.2359 | 0.00e+00 | 0.3417 | 3.44e-18 | 0.3607 | 1.14e-04 | Cubic | 0.3607 | Cubic (p_quad=0.0000 |
| PS | OAE at 5273 Hz | 511 | 0.1865 | 0.00e+00 | 0.1934 | 3.76e-02 | 0.2192 | 4.95e-05 | Cubic | 0.2192 | Cubic (p_quad=0.0376 |
| PS | Wave I amplitude - Eclipse | 134 | 0.0452 | 1.36e-02 | 0.0479 | 5.41e-01 | 0.0489 | 7.26e-01 | Linear | 0.0452 | Linear (p_quad=0.5412) |
| PS | Wave I latency | 317 | 0.1010 | 7.22e-09 | 0.1154 | 2.43e-02 | 0.1161 | 6.23e-01 | Quadratic | 0.1154 | Quadratic (p_quad=0.0243 |
| PS | Wave III latency | 315 | 0.0306 | 1.82e-03 | 0.0637 | 1.01e-03 | 0.1118 | 5.15e-05 | Cubic | 0.1118 | Cubic (p_quad=0.0010 |
| PS | Wave V latency | 314 | 0.0583 | 1.52e-05 | 0.0583 | 9.86e-01 | 0.0585 | 8.11e-01 | Linear | 0.0583 | Linear (p_quad=0.9859) |

### Table S8: Hierarchical model progression for combined-device latency and Eclipse-only amplitude models

| a) Model Phase ABR Latency | Outcome | Observations (n) | Parameters_including_intercept | R2 | Adjusted_R2 | Delta_R2 | Nested_model_p | AIC |
|---|---|---|---|---|---|---|---|---|
| Phase 0: age + gender | W S | 298 | 4 | 0.22663348294774 | 0.218741987875778 | | | 2043.74976584061 |
| Phase 1: + hearing measures | W S | 298 | 12 | 0.290578115343971 | 0.263292658241816 | 0.0639446323962317 | 0.00159207023241124 | 2034.0315631299 |
| Phase 2: + Wave I/III/V latencies | W S | 298 | 17 | 0.339901425392314 | 0.302315741428887 | 0.0493233100483422 | 0.00107114747590422 | 2022.55732193642 |
| Phase 3: + age x Wave I latency | W S | 298 | 18 | 0.361944108096233 | 0.323205000373504 | 0.0220426827039192 | 0.00206279305194847 | 2014.43626003783 |
| Phase 0: age + gender | P S | 298 | 4 | 0.240109710198044 | 0.232355727649044 | | | 1731.48380660497 |
| Phase 1: + hearing measures | P S | 298 | 12 | 0.332375315216958 | 0.306697442725302 | 0.0922656050189143 | 9.69284818224119e-06 | 1708.90833215963 |
| Phase 2: + Wave I/III/V latencies | P S | 298 | 17 | 0.399449392341266 | 0.365254339947886 | 0.0670740771243082 | 1.53683606406108e-05 | 1687.35631486216 |
| Phase 3: + age x Wave I latency | P S | 298 | 18 | 0.415442586547204 | 0.379951600730427 | 0.0159931942059377 | 0.00602008196173984 | 1681.31272534241 |
| b) Model Phase ABR Latency | Outcome | Observations (n) | Parameters_including_intercept | R2 | Adjusted_R2 | Delta_R2 | Nested_model_p | AIC |
| Phase 0: age + gender | W S | 122 | 4 | 0.112714520689988 | 0.0901564152838019 | | | 852.676649180219 |
| Phase 1: + hearing measures | W S | 122 | 12 | 0.288901062125831 | 0.217791168338414 | 0.176186541435842 | 0.00155698722531094 | 841.671314117992 |
| Phase 2: + Wave I/III/V latencies | W S | 122 | 17 | 0.34469983900848 | 0.244844576381201 | 0.0557987768826497 | 0.121580074929064 | 841.701695971443 |
| Phase 3: + age x Wave I latency | W S | 122 | 18 | 0.360884290235608 | 0.256413453062582 | 0.0161844512271273 | 0.107650985155903 | 840.650735307654 |
| Phase 4: + Wave I amplitude | W S | 122 | 19 | 0.362874294100681 | 0.251531937729926 | 0.00199000386507314 | 0.571815266076335 | 842.270273327156 |
| Phase 5: + age x Wave I amplitude | W S | 122 | 20 | 0.363086342985999 | 0.244445563738293 | 0.000212048885318283 | 0.854159813458941 | 844.229662393145 |
| Phase 0: age + gender | P S | 122 | 4 | 0.109445516172115 | 0.0868043004815756 | | | 727.851654907315 |
| Phase 1: + hearing measures | P S | 122 | 12 | 0.371102052421812 | 0.308212257663993 | 0.261656536249697 | 4.37589254877571e-06 | 701.410869986525 |
| Phase 2: + Wave I/III/V latencies | P S | 122 | 17 | 0.435456819033775 | 0.349431191457969 | 0.064354766611963 | 0.042409752861421 | 698.240710957773 |
| Phase 3: + age x Wave I latency | P S | 122 | 18 | 0.453786231317002 | 0.364501288359205 | 0.0183294122832277 | 0.064557361440368 | 696.213923082898 |
| Phase 4: + Wave I amplitude | P S | 122 | 19 | 0.457005958528318 | 0.362113795941033 | 0.00321972721131519 | 0.436299429684832 | 697.492650502493 |
| Phase 5: + age x Wave I amplitude | P S | 122 | 20 | 0.457875834651811 | 0.356891921498718 | 0.000869876123493817 | 0.686650482511527 | 699.297049858927 |

Note. Panel a presents sequential models for ABR latency using eligible Eclipse and ChartR observations (n = 298); panel b presents a separate Eclipse-only sequence that adds Wave I amplitude and the age × Wave I amplitude interaction (n = 122). Nested_model_p values are conventional partial F-test p values from comparisons of adjacent nested ordinary least-squares models fitted to the same observations; they are not based on HC3 heteroscedasticity-robust covariance estimates and are not adjusted for multiple testing. $\Delta R^2$ is reported only for valid within-sample nested comparisons. WS, word score; PS, phoneme score; ABR, auditory brainstem response; $R^2$, coefficient of determination; AIC, Akaike information criterion.

#### Table S9: Combined-device latency model coefficients with HC3-robust inference (n = 298)

| Variable | WS estimate | WS robust_se | WS_t | WS_CI_Lower | WS_CI_Upper | WS robust_p |
|---|---|---|---|---|---|---|
| (Intercept) | 78.00864730937 | 0.89296425495076 | 87.3592048918817 | 76.2508717302512 | 79.7664228884887 | 3.39331903467868e-205 |
| Age_c | -0.0196579197351512 | 0.0603067890638954 | -0.325965285837445 | -0.13837017555487 | 0.0990543360845681 | 0.744694100078894 |
| Age_c2 | -0.0100940974149217 | 0.00397507164501143 | -2.53934980708825 | -0.0179189166326111 | -0.00226927819723238 | 0.0116470872555488 |
| GenderWoman | 0.41858953808955 | 0.942070736087093 | 0.444329201677752 | -1.4358507987049 | 2.273029874884 | 0.657147814322839 |
| PTA4_c | 0.0342408338592581 | 0.0926898891461226 | 0.369412825656513 | -0.148216663928557 | 0.216698331647073 | 0.712099255339692 |
| PTA4_c2 | -0.00324240199566044 | 0.00833379614213419 | -0.389066631863891 | -0.019647250519866 | 0.0131624465285451 | 0.697522570411127 |
| PTAHF_c | -0.0700611065042827 | 0.0545489638313976 | -1.28437098678593 | -0.177439241695538 | 0.0373170286869728 | 0.200073814901835 |
| PTAHF_c2 | -0.000117948961033499 | 0.00138570662658447 | -0.0851182773977373 | -0.00284567434816838 | 0.00260977642610138 | 0.932228188827781 |
| PTAHF_c3 | -5.15343772759089e-06 | 4.54246035625431e-05 | -0.113450362213846 | -9.45705206519133e-05 | 8.42636451967315e-05 | 0.909754814155132 |
| OAE_c | -0.0035466518418342 | 0.113033609243005 | -0.0313769671302755 | -0.226050203182711 | 0.218956899499042 | 0.974991263488068 |
| OAE_c2 | 0.000398624560728681 | 0.00664472102620109 | 0.0599911657926416 | -0.0126813260477725 | 0.0134785751692298 | 0.952205465670148 |
| OAE_c3 | 0.000277170432476291 | 0.000447081533342402 | 0.619955001057978 | -0.000602897271048486 | 0.00115723813600106 | 0.535791605528757 |
| W1Lat_c | -9.92456245494509 | 4.81321347636328 | -2.06194105116729 | -19.3992408185298 | -0.449884091360353 | 0.0401359958398797 |
| W3Lat_c | -7.17889825928919 | 5.07011045368753 | -1.41592541718059 | -17.1592712580022 | 2.80147473942389 | 0.157908746193659 |
| W3Lat_c2 | 9.43715021019851 | 10.2835708457063 | 0.917691952707146 | -10.8057761986717 | 29.6800766190687 | 0.359569946768356 |
| W3Lat_c3 | 81.782688214276 | 35.9227095044664 | 2.27662916696491 | 11.0698224386922 | 152.495553989859 | 0.0235631366740031 |
| W5Lat_c | -0.672797827539072 | 1.48270132461241 | -0.453764906236221 | -3.59145461036225 | 2.2458589552841 | 0.650349542809212 |
| Age_x_W1Lat | -0.788436929780637 | 0.353514044332077 | -2.2302846023283 | -1.48431960431914 | -0.0925542552421299 | 0.0265211112815733 |
| Variable | PS estimate | PS robust_se | PS_t | PS_CI_Lower | PS_CI_Upper | PS robust_p |
| (Intercept) | 91.5004712135308 | 0.514388377598256 | 177.88207354287 | 90.4879118380443 | 92.5130305890173 | 5.1206247710098e-290 |
| Age_c | -0.016009807043851 | 0.032715770258423 | -0.489360541334926 | -0.0804099011052711 | 0.048390287017569 | 0.624969598843797 |
| Age_c2 | -0.00413775610446394 | 0.00259956122220501 | -1.59171327419409 | -0.00925492088175727 | 0.000979408672829381 | 0.112577450120739 |
| GenderWoman | -0.187150320050673 | 0.570634319503945 | -0.327968917490564 | -1.31042828540033 | 0.93612764529899 | 0.743180270489087 |
| PTA4_c | 0.0152125016206368 | 0.0497883192878975 | 0.305543586090376 | -0.0827944357057156 | 0.113219438946989 | 0.760179205165481 |
| PTA4_c2 | -0.00480812769038536 | 0.00630621318188238 | -0.762442935516199 | -0.0172217349163999 | 0.0076054795356292 | 0.446437493131657 |
| PTAHF_c | -0.0291504671557075 | 0.0308165959298974 | -0.94593404222906 | -0.0898120887792185 | 0.0315111544678034 | 0.344997814629194 |
| PTAHF_c2 | -0.00112872527330468 | 0.000936943401572774 | -1.20468885464156 | -0.00297307259311074 | 0.000715622046501384 | 0.229340697722191 |
| PTAHF_c3 | -9.08917304859001e-06 | 3.13479744303251e-05 | -0.28994450881641 | -7.07967986371443e-05 | 5.26184525399643e-05 | 0.772073290791071 |
| OAE_c | 0.0425495161063241 | 0.0663990136858628 | 0.640815484212282 | -0.0881551163439587 | 0.173254148556607 | 0.522166685217647 |
| OAE_c2 | 0.000897283030795938 | 0.00372937075221045 | 0.240599042147822 | -0.0064438807362539 | 0.00823844679784577 | 0.810041961335362 |
| OAE_c3 | 9.57844706737917e-05 | 0.000278884750914523 | 0.343455389223304 | -0.000453192492097455 | 0.000644761433445038 | 0.73151364131577 |
| W1Lat_c | -4.93175432351891 | 3.08958056235962 | -1.5962536739137 | -11.0135086962458 | 1.15000004920803 | 0.111560149028577 |
| W3Lat_c | -6.38020848778096 | 3.319691869446 | -1.92192791942636 | -12.9149306403859 | 0.154513664824061 | 0.0556295522502739 |

| Variable | WS estimate | WS robust_se | WS_t | WS_CI_Lower | WS_CI_Upper | WS robust_p |
|---|---|---|---|---|---|---|
| W3Lat_c2 | 3.03437298517678 | 7.30923282909717 | 0.415142472011195 | -11.353650833076 | 17.4223968034295 | 0.678355535140073 |
| W3Lat_c3 | 67.4248720803471 | 30.3220964007687 | 2.2236217176144 | 7.73665918105898 | 127.113084979635 | 0.0269718536591355 |
| W5Lat_c | -0.334991140719891 | 0.759945535025307 | -0.440809407096175 | -1.83092302771514 | 1.16094074627536 | 0.659691130494953 |
| Age_x_W1Lat | -0.401214837310632 | 0.225005295177674 | -1.78313509019338 | -0.844131573163096 | 0.0417018985418309 | 0.0756475021843984 |

Note. Latency models include eligible Eclipse and ChartR observations (n = 298) and exclude Wave I amplitude. Standard errors, t statistics, 95% confidence intervals, and p values are based on the HC3 heteroscedasticity-robust covariance estimator. HC3 inference is robust to heteroscedasticity and leverage but does not constitute adjustment for multiple testing. Continuous predictors were mean-centred before construction of polynomial and interaction terms. WS, word score; PS, phoneme score; PTA4, pure-tone average at 0.5, 1, 2, and 4 kHz; PTAHF, extended-high-frequency pure-tone average; OAE, otoacoustic emission; W1Lat, Wave I latency; W3Lat, Wave III latency; W5Lat, Wave V latency; c, centred; c2, squared centred term; c3, cubed centred term; Age_x_W1Lat, age × Wave I latency interaction.

#### Table S10: Eclipse-only Wave I amplitude model coefficients with HC3-robust inference (n = 122)

| Variable | WS estimate | WS robust_se | WS_t | WS_CI_Lower | WS_CI_Upper | WS robust_p |
|---|---|---|---|---|---|---|
| (Intercept) | 81.2062630709419 | 1.72460876468195 | 47.0867739593782 | 77.7855097633192 | 84.6270163785645 | 5.16240439677695e-71 |
| Age_c | 0.0809897076115588 | 0.134257490326078 | 0.603241632290719 | -0.185309387876768 | 0.347288803099886 | 0.547687015222862 |
| Age_c2 | -0.00406196845544828 | 0.00870173992947413 | -0.466799569783713 | -0.0213218283468075 | 0.0131978914359109 | 0.641638827418368 |
| GenderWoman | -0.590440029616042 | 2.06898113509724 | -0.285377193440814 | -4.69425430113747 | 3.51337424190539 | 0.775933866252133 |
| PTA4_c | -0.110174448762973 | 0.196001800433821 | -0.56210937103189 | -0.498943090593246 | 0.278594193067299 | 0.57527469392148 |
| PTA4_c2 | -0.0166515249207165 | 0.0208159278549903 | -0.799941517702972 | -0.0579398191236769 | 0.0246367692822437 | 0.425603784062962 |
| PTAHF_c | -0.029436657343084 | 0.124434055335374 | -0.236564317250181 | -0.276251016104549 | 0.217377701418381 | 0.81346922668974 |
| PTAHF_c2 | -0.00412691266692289 | 0.00297761388300815 | -1.38597979089003 | -0.0100329956857005 | 0.00177917035185477 | 0.168775705429612 |
| PTAHF_c3 | 4.15183037251205e-05 | 0.000115724314874419 | 0.358769060505347 | -0.000188020326128728 | 0.000271056933578969 | 0.720509397166572 |
| OAE_c | 0.130671377029018 | 0.234256966826774 | 0.557812127421789 | -0.33397620595721 | 0.595318960015247 | 0.578194913338662 |
| OAE_c2 | 0.0224828964871521 | 0.0157776003286742 | 1.42498833908802 | -0.00881189895627328 | 0.0537776919305775 | 0.157213288215196 |
| OAE_c3 | 0.00100905084467998 | 0.0014758007764294 | 0.683731070477758 | -0.00191819299795116 | 0.00393629468731113 | 0.49569534360913 |
| W1Lat_c | -4.98801907783254 | 13.769312540687 | -0.362256217446833 | -32.2993852159557 | 22.3233470602906 | 0.717910040381402 |
| W3Lat_c | -11.3977811529065 | 10.6751104506101 | -1.06769678924072 | -32.5718121163267 | 9.77624981051368 | 0.288177793955054 |
| W3Lat_c2 | -38.2855590131845 | 23.7810821262277 | -1.60991660555934 | -85.4552226540516 | 8.88410462768251 | 0.110506677636285 |
| W3Lat_c3 | 99.3232495748513 | 93.432296568197 | 1.0630504999131 | -85.9992676648082 | 284.64576681451 | 0.290269369126226 |
| W5Lat_c | -2.08643722790865 | 2.7609127069141 | -0.755705612380871 | -7.56269449137879 | 3.38982003556148 | 0.45156737541249 |
| Age_x_W1Lat | -0.828204087871787 | 1.09189620960699 | -0.758500744470834 | -2.99397504247004 | 1.33756686672646 | 0.449900360314188 |
| W1Amp_c | 2.35186491031505 | 5.12508107668481 | 0.458893210687775 | -7.81370910503961 | 12.5174389256697 | 0.647287230976024 |
| Age_x_W1Amp | -0.110332944670912 | 0.643148552928448 | -0.17155126007597 | -1.38601504995606 | 1.16534916061424 | 0.864130211391081 |
| Variable | PS estimate | PS robust_se | PS_t | PS_CI_Lower | PS_CI_Upper | PS robust_p |
| (Intercept) | 93.3456432185728 | 1.01050919093619 | 92.3748581960861 | 91.3413030296167 | 95.349983407529 | 3.87294851378254e-100 |
| Age_c | 0.0370370740258346 | 0.0724758214090143 | 0.511026619716628 | -0.106718374099395 | 0.180792522151064 | 0.610436777583654 |
| Age_c2 | 0.00102889977737045 | 0.00522545502124573 | 0.196901470434084 | -0.00933576548110405 | 0.0113935650358449 | 0.844296453044877 |
| GenderWoman | -1.02466292690758 | 1.20743575402585 | -0.848627285957978 | -3.41960602003717 | 1.37028016622199 | 0.398075919461894 |
| PTA4_c | -0.0893335740263508 | 0.118440635392451 | -0.75424767631772 | -0.324260012748453 | 0.145592864695751 | 0.452438292615645 |
| PTA4_c2 | -0.017929752384793 | 0.0198072134585939 | -0.905213265978799 | -0.057217266365257 | 0.0213577615956709 | 0.36748563607964 |
| PTAHF_c | -0.0125760373818134 | 0.0692111389994081 | -0.181705395455505 | -0.149856003426875 | 0.124703928663249 | 0.856174538433853 |
| PTAHF_c2 | -0.00304616985836796 | 0.00186219562692133 | -1.6357947652385 | -0.00673982605488295 | 0.000647486338147036 | 0.104964768876461 |
| PTAHF_c3 | 2.26897336042133e-05 | 6.68007216068366e-05 | 0.339663001513008 | -0.000109809180971526 | 0.000155188648179952 | 0.734808729652554 |
| OAE_c | 0.0822456435372386 | 0.106132725943138 | 0.774931980747415 | -0.128268115149329 | 0.292759402223806 | 0.440172628301629 |
| OAE_c2 | 0.0148187468310766 | 0.00921305143858491 | 1.60845154614189 | -0.0034552970142524 | 0.0330927906764057 | 0.110827311210339 |
| OAE_c3 | 0.000454951032055915 | 0.00052859480165859 | 0.86068011003589 | -0.000593514250734885 | 0.00150341631484671 | 0.391432861687806 |

| Variable | WS estimate | WS robust_se | WS_t | WS_CI_Lower | WS_CI_Upper | WS robust_p |
|---|---|---|---|---|---|---|
| W1Lat_c | -1.18805530888638 | 9.88877808258163 | -0.120141770698551 | -20.8023997486675 | 18.4262891308947 | 0.904607173929281 |
| W3Lat_c | -7.7419053011097 | 5.71474652865474 | -1.35472417932981 | -19.077077944585 | 3.59326734236565 | 0.178498543240686 |
| W3Lat_c2 | -25.0722923533911 | 15.0397578780461 | -1.66706755233006 | -54.9035807944293 | 4.75899608764703 | 0.0985687154554393 |
| W3Lat_c3 | 54.63989816324 | 51.6913425816848 | 1.05704157474524 | -47.8896347562813 | 157.169431082761 | 0.292989688018685 |
| W5Lat_c | -1.17660266933728 | 1.46039209487601 | -0.805675868464068 | -4.07328346516656 | 1.72007812649198 | 0.422304038644028 |
| Age_x_W1Lat | -0.48415390020405 | 0.965213338376389 | -0.501602993819437 | -2.39864998037526 | 1.43034217996716 | 0.617027765753153 |
| W1Amp_c | 2.2791519196478 | 3.14191573273781 | 0.725401988315514 | -3.95282303904176 | 8.51112687833737 | 0.469866470415454 |
| Age_x_W1Amp | 0.13373402690839 | 0.422610012374223 | 0.316447843147568 | -0.704510928857082 | 0.971978982673862 | 0.752309307830085 |

Note. Models are restricted to Eclipse observations with valid Wave I amplitude (n = 122). Standard errors, t statistics, 95% confidence intervals, and p values are based on the HC3 heteroscedasticity-robust covariance estimator. HC3 inference is robust to heteroscedasticity and leverage but does not constitute adjustment for multiple testing. Continuous predictors were mean-centred before construction of polynomial and interaction terms. WS, word score; PS, phoneme score; PTA4, pure-tone average at 0.5, 1, 2, and 4 kHz; PTAHF, extended-high-frequency pure-tone average; OAE, otoacoustic emission; W1Lat, Wave I latency; W3Lat, Wave III latency; W5Lat, Wave V latency; W1Amp, Wave I amplitude; c, centred; c2, squared centred term; c3, cubed centred term; Age_x_W1Lat, age × Wave I latency interaction; Age_x_W1Amp, age × Wave I amplitude interaction.

#### Table S11: Sensitivity analyses of the final multivariable models for word and phoneme scores (WS, PS): heteroscedasticity-robust (HC3) versus participant-clustered (CR3) inference, and adjustment for recording device

| Word score (WS): model | Term | β | HC3 SE | HC3 95% CI lower | HC3 95% CI upper | HC3 p | CR3 SE | CR3 df (Satterthwaite) | CR3 95% CI lower | CR3 95% CI upper | CR3 p |
|---|---|---|---|---|---|---|---|---|---|---|---|
| Latency model (n = 298) | Age$^2$ (Age_c2) | -0.01009 | 0.003975 | -0.01792 | -0.002269 | 0.01165 | 0.003674 | 32.39 | -0.01757 | -0.002614 | 0.009728 |
| Latency model (n = 298) | Wave I latency (W1Lat_c) | -9.925 | 4.813 | -19.4 | -0.4499 | 0.04014 | 4.879 | 56.05 | -19.7 | -0.1519 | 0.04666 |
| Latency model (n = 298) | Age x Wave I latency (Age_x_W1Lat) | -0.7884 | 0.3535 | -1.484 | -0.09255 | 0.02652 | 0.347 | 19.55 | -1.513 | -0.06346 | 0.03455 |
| Latency model (n = 298) | Wave III latency$^3$ (W3Lat_c3) | 81.78 | 35.92 | 11.07 | 152.5 | 0.02356 | 36.74 | 10.55 | 0.4977 | 163.1 | 0.04884 |
| Latency model (n = 298) | Wave V latency (W5Lat_c) | -0.6728 | 1.483 | -3.591 | 2.246 | 0.6503 | 1.532 | 68.98 | -3.73 | 2.384 | 0.662 |
| Latency model (n = 298) | Gender, women vs men (GenderWoman) | 0.4186 | 0.9421 | -1.436 | 2.273 | 0.6571 | 1.009 | 81.66 | -1.588 | 2.425 | 0.6792 |
| Latency model + device | Age$^2$ (Age_c2) | -0.01029 | 0.004007 | -0.01818 | -0.002404 | 0.01074 | 0.003656 | 30.8 | -0.01775 | -0.002834 | 0.008422 |
| Latency model + device | Wave I latency (W1Lat_c) | -10.92 | 5.812 | -22.36 | 0.523 | 0.06135 | 5.895 | 43.41 | -22.8 | 0.9663 | 0.0708 |
| Latency model + device | Age x Wave I latency (Age_x_W1Lat) | -0.7899 | 0.3544 | -1.487 | -0.09234 | 0.0266 | 0.3482 | 19.41 | -1.518 | -0.06217 | 0.03487 |
| Latency model + device | Wave III latency$^3$ (W3Lat_c3) | 83.79 | 38.21 | 8.583 | 159 | 0.02912 | 38.85 | 10.36 | -2.367 | 170 | 0.05549 |
| Latency model + device | Wave V latency (W5Lat_c) | -0.734 | 1.506 | -3.698 | 2.23 | 0.6263 | 1.559 | 67.99 | -3.845 | 2.378 | 0.6394 |
| Latency model + device | Gender, women vs men (GenderWoman) | 0.3541 | 0.9523 | -1.521 | 2.229 | 0.7103 | 1.021 | 76.89 | -1.678 | 2.386 | 0.7296 |
| Latency model + device | Device, Eclipse vs Chartr (DeviceEclipse) | -0.5263 | 1.637 | -3.749 | 2.697 | 0.7481 | 1.666 | 53.41 | -3.867 | 2.815 | 0.7533 |
| Eclipse amplitude model (n = 122) | Age$^2$ (Age_c2) | -0.004062 | 0.008702 | -0.02132 | 0.0132 | 0.6416 | 0.007766 | 11.12 | -0.02113 | 0.01301 | 0.6112 |
| Eclipse amplitude model (n = 122) | Wave I latency (W1Lat_c) | -4.988 | 13.77 | -32.3 | 22.32 | 0.7179 | 14.24 | 16.14 | -35.15 | 25.17 | 0.7306 |
| Eclipse amplitude model (n = 122) | Age x Wave I latency (Age_x_W1Lat) | -0.8282 | 1.092 | -2.994 | 1.338 | 0.4499 | 1.115 | 4.172 | -3.873 | 2.217 | 0.4971 |
| Eclipse amplitude model (n = 122) | Wave III latency$^3$ (W3Lat_c3) | 99.32 | 93.43 | -86 | 284.6 | 0.2903 | 87.46 | 12.41 | -90.54 | 289.2 | 0.2776 |
| Eclipse amplitude model (n = 122) | Wave V latency (W5Lat_c) | -2.086 | 2.761 | -7.563 | 3.39 | 0.4516 | 2.971 | 30.46 | -8.151 | 3.978 | 0.4879 |
| Eclipse amplitude model (n = 122) | Gender, women vs men (GenderWoman) | -0.5904 | 2.069 | -4.694 | 3.513 | 0.7759 | 2.276 | 26.97 | -5.26 | 4.079 | 0.7973 |
| Eclipse amplitude model (n = 122) | Wave I amplitude (W1Amp_c) | 2.352 | 5.125 | -7.814 | 12.52 | 0.6473 | 5.036 | 18.17 | -8.221 | 12.92 | 0.646 |
| Eclipse amplitude model (n = 122) | Age x Wave I amplitude (Age_x_W1Amp) | -0.1103 | 0.6431 | -1.386 | 1.165 | 0.8641 | 0.6281 | 16.59 | -1.438 | 1.217 | 0.8627 |
| Phoneme score (PS): model | Term | β | HC3 SE | HC3 95% CI lower | HC3 95% CI upper | HC3 p | CR3 SE | CR3 df (Satterthwaite) | CR3 95% CI lower | CR3 95% CI upper | CR3 p |
| Latency model (n = 298) | Age$^2$ (Age_c2) | -0.004138 | 0.0026 | -0.009255 | 0.0009794 | 0.1126 | 0.00258 | 32.39 | -0.00939 | 0.001114 | 0.1184 |
| Latency model (n = 298) | Wave I latency (W1Lat_c) | -4.932 | 3.09 | -11.01 | 1.15 | 0.1116 | 3.174 | 56.05 | -11.29 | 1.427 | 0.1259 |
| Latency model (n = 298) | Age x Wave I latency (Age_x_W1Lat) | -0.4012 | 0.225 | -0.8441 | 0.0417 | 0.07565 | 0.2293 | 19.55 | -0.8802 | 0.07774 | 0.0958 |
| Latency model (n = 298) | Wave III latency$^3$ (W3Lat_c3) | 67.42 | 30.32 | 7.737 | 127.1 | 0.02697 | 30.66 | 10.55 | -0.414 | 135.3 | 0.05118 |
| Latency model (n = 298) | Wave V latency (W5Lat_c) | -0.335 | 0.7599 | -1.831 | 1.161 | 0.6597 | 0.7955 | 68.98 | -1.922 | 1.252 | 0.675 |
| Latency model (n = 298) | Gender, women vs men (GenderWoman) | -0.1872 | 0.5706 | -1.31 | 0.9361 | 0.7432 | 0.628 | 81.66 | -1.436 | 1.062 | 0.7664 |
| Latency model + device | Age$^2$ (Age_c2) | -0.004261 | 0.002564 | -0.009308 | 0.0007855 | 0.09761 | 0.00252 | 30.8 | -0.009402 | 0.0008797 | 0.1009 |
| Latency model + device | Wave I latency (W1Lat_c) | -5.554 | 3.942 | -13.31 | 2.206 | 0.16 | 3.993 | 43.41 | -13.6 | 2.496 | 0.1713 |
| Latency model + device | Age x Wave I latency (Age_x_W1Lat) | -0.4021 | 0.2262 | -0.8474 | 0.04314 | 0.07653 | 0.2306 | 19.41 | -0.8842 | 0.07992 | 0.09706 |
| Latency model + device | Wave III latency$^3$ (W3Lat_c3) | 68.68 | 32.19 | 5.327 | 132 | 0.03371 | 32.62 | 10.36 | -3.652 | 141 | 0.06054 |
| Latency model + device | Wave V latency (W5Lat_c) | -0.3733 | 0.778 | -1.905 | 1.158 | 0.6317 | 0.8136 | 67.99 | -1.997 | 1.25 | 0.6478 |
| Latency model + device | Gender, women vs men (GenderWoman) | -0.2276 | 0.5973 | -1.403 | 0.9483 | 0.7035 | 0.6561 | 76.89 | -1.534 | 1.079 | 0.7296 |

| Word score (WS): model | Term | β | HC3 SE | HC3 95% CI lower | HC3 95% CI upper | HC3 p | CR3 SE | CR3 df (Satterthwaite) | CR3 95% CI lower | CR3 95% CI upper | CR3 p |
|---|---|---|---|---|---|---|---|---|---|---|---|
| Latency model + device | Device, Eclipse vs Chartr (DeviceEclipse) | -0.3297 | 1.157 | -2.607 | 1.948 | 0.7759 | 1.196 | 53.41 | -2.729 | 2.069 | 0.7839 |
| Eclipse amplitude model (n = 122) | Age² (Age_c2) | 0.001029 | 0.005225 | -0.009336 | 0.01139 | 0.8443 | 0.005346 | 11.12 | -0.01072 | 0.01278 | 0.8509 |
| Eclipse amplitude model (n = 122) | Wave I latency (W1Lat_c) | -1.188 | 9.889 | -20.8 | 18.43 | 0.9046 | 10.16 | 16.14 | -22.72 | 20.34 | 0.9084 |
| Eclipse amplitude model (n = 122) | Age x Wave I latency (Age_x_W1Lat) | -0.4842 | 0.9652 | -2.399 | 1.43 | 0.617 | 0.9788 | 4.172 | -3.158 | 2.19 | 0.6458 |
| Eclipse amplitude model (n = 122) | Wave III latency³ (W3Lat_c3) | 54.64 | 51.69 | -47.89 | 157.2 | 0.293 | 51.62 | 12.41 | -57.43 | 166.7 | 0.31 |
| Eclipse amplitude model (n = 122) | Wave V latency (W5Lat_c) | -1.177 | 1.46 | -4.073 | 1.72 | 0.4223 | 1.549 | 30.46 | -4.339 | 1.986 | 0.4535 |
| Eclipse amplitude model (n = 122) | Gender, women vs men (GenderWoman) | -1.025 | 1.207 | -3.42 | 1.37 | 0.3981 | 1.329 | 26.97 | -3.752 | 1.703 | 0.4475 |
| Eclipse amplitude model (n = 122) | Wave I amplitude (W1Amp_c) | 2.279 | 3.142 | -3.953 | 8.511 | 0.4699 | 3.303 | 18.17 | -4.656 | 9.214 | 0.4989 |
| Eclipse amplitude model (n = 122) | Age x Wave I amplitude (Age_x_W1Amp) | 0.1337 | 0.4226 | -0.7045 | 0.972 | 0.7523 | 0.4646 | 16.59 | -0.8484 | 1.116 | 0.777 |

Note. Clusters are participants (163 in the latency models; 68 in the Eclipse-only model), as both ears of a participant were entered when available. HC3, heteroscedasticity-consistent standard errors treating ears as independent (identical to Tables S9 and S10). CR3, jackknife-type cluster-robust standard errors (Bell & McCaffrey, 2002) with Satterthwaite small-sample degrees of freedom, computed with the clubSandwich R package (Pustejovsky & Tipton, 2018). "Latency model + device" adds recording device (Eclipse vs Chartr) as a covariate to the combined-device latency model (n = 298 ears). All models also include PTA4 (linear, quadratic), eHF PTA, OAE and Wave III latency (linear, quadratic, cubic) terms, which are not shown. Continuous predictors were mean-centred before construction of polynomial and interaction terms. Low Satterthwaite degrees of freedom (e.g., < 15) indicate that inference for that term rests on few effectively informative participants and should be interpreted cautiously. No adjustment for multiple testing was applied. WS, word score; PS, phoneme score; β, unstandardized coefficient; SE, standard error; CI, confidence interval.

#### Table S12: Combined-device latency model predictions with HC3-robust uncertainty

| Age Group | W1.Latency Level | W1.Lat (ms) | WS Predicted | WS SE | WS 95% CI [L] | WS 95% CI [U] |
|---|---|---|---|---|---|---|
| 45 | Minimum | 1 | 81.7124466107914 | 2.0246128634157 | 77.7270518665437 | 85.697841355039 |
| 45 | Q1 | 1.2667 | 79.445475204511 | 1.03506818327412 | 77.4079719503831 | 81.4829784586388 |
| 45 | Median | 1.4 | 78.3119895056214 | 0.832063328751981 | 76.6740957249335 | 79.9498832863093 |
| 45 | Q3 | 1.48 | 77.6318980845873 | 0.909970404652512 | 75.8406463674543 | 79.4231498017204 |
| 45 | Maximum | 1.83 | 74.6564981175636 | 2.15037101093453 | 70.4235519183073 | 78.8894443168199 |
| 55 | Minimum | 1 | 83.9153892392034 | 2.64570673552106 | 78.7073882879612 | 89.1233901904456 |
| 55 | Q1 | 1.2667 | 79.5459193508798 | 1.23258312401519 | 77.119613367616 | 81.9722253341437 |
| 55 | Median | 1.4 | 77.3611844149108 | 1.00903462334758 | 75.3749275075686 | 79.347441322253 |
| 55 | Q3 | 1.48 | 76.0503434500523 | 1.19750151558002 | 73.6930946482396 | 78.407592251865 |
| 55 | Maximum | 1.83 | 70.3154142287963 | 3.09385998441103 | 64.2252359314464 | 76.4055925261462 |
| 65 | Minimum | 1 | 84.099512384631 | 4.15348885213807 | 75.9234838012329 | 92.2755409680291 |
| 65 | Q1 | 1.2667 | 77.6275440142643 | 2.22745110658558 | 73.2428677664418 | 82.0122202620868 |
| 65 | Median | 1.4 | 74.3915598412159 | 1.86897913898015 | 70.7125258035513 | 78.0705938788805 |
| 65 | Q3 | 1.48 | 72.4499693325329 | 2.02694179253446 | 68.4599901553611 | 76.4399485097046 |
| 65 | Maximum | 1.83 | 63.9555108570447 | 4.47184961669488 | 55.1527978636213 | 72.7582238504681 |
| 75 | Minimum | 1 | 82.2648160470743 | 6.42371644024273 | 69.6199068883479 | 94.9097252058007 |
| 75 | Q1 | 1.2667 | 73.6903491946644 | 4.12371140834739 | 65.5729366904808 | 81.8077616988481 |
| 75 | Median | 1.4 | 69.4031157845367 | 3.64997849266546 | 62.2182334970931 | 76.5879980719802 |
| 75 | Q3 | 1.48 | 66.8307757320291 | 3.71695633006413 | 59.5140494135744 | 74.1475020504838 |
| 75 | Maximum | 1.83 | 55.5767880023087 | 6.30956600884295 | 43.1565808286259 | 67.9969951759915 |
| Age Group | W1.Latency Level | W1.Lat (ms) | PS Predicted | PS SE | PS 95% CI [L] | PS 95% CI [U] |
| 45 | Minimum | 1 | 92.9533832451944 | 1.30592847173469 | 90.382698961961 | 95.5240675284277 |
| 45 | Q1 | 1.2667 | 91.8314063257288 | 0.647463506308937 | 90.5568922189763 | 93.1059204324812 |
| 45 | Median | 1.4 | 91.2704178680997 | 0.474313388973514 | 90.3367450056483 | 92.2040907305511 |
| 45 | Q3 | 1.48 | 90.9338247926808 | 0.502070395540151 | 89.9455130261279 | 91.9221365592337 |
| 45 | Maximum | 1.83 | 89.461230087723 | 1.27958550194888 | 86.9424012160744 | 91.9800589593715 |
| 55 | Minimum | 1 | 94.078158873754 | 1.8174153719007 | 90.5006266935681 | 97.6556910539398 |
| 55 | Q1 | 1.2667 | 91.8862757201227 | 0.81048660168994 | 90.2908551332907 | 93.4816963069546 |
| 55 | Median | 1.4 | 90.7903341474168 | 0.580935032584594 | 89.6467795116402 | 91.9338887831934 |
| 55 | Q3 | 1.48 | 90.1327692021493 | 0.6833830176692 | 88.7875485250802 | 91.4779898792185 |
| 55 | Maximum | 1.83 | 87.2559225666043 | 1.96618229826259 | 83.3855467830047 | 91.126298350204 |
| 65 | Minimum | 1 | 94.3753832814208 | 2.78106496261694 | 88.9009333825272 | 99.8498331803144 |
| 65 | Q1 | 1.2667 | 91.1135938936238 | 1.43653533376804 | 88.2858135942008 | 93.9413741930467 |
| 65 | Median | 1.4 | 89.4826992058411 | 1.1518396926454 | 87.2153344467631 | 91.7500639649191 |
| 65 | Q3 | 1.48 | 88.5041623907251 | 1.24743644441491 | 86.0486180547247 | 90.9597067267256 |
| 65 | Maximum | 1.83 | 84.2230638245929 | 2.91223468228732 | 78.4904099397299 | 89.9557177094559 |
| 75 | Minimum | 1 | 93.8450564681949 | 4.20992383718581 | 85.5579371687072 | 102.132175767682 |
| 75 | Q1 | 1.2667 | 89.5133608462321 | 2.64971663170935 | 84.2974665246164 | 94.7292551678477 |
| 75 | Median | 1.4 | 87.3475130433726 | 2.32444224680729 | 82.7719124033055 | 91.9231136834397 |
| 75 | Q3 | 1.48 | 86.0480043584082 | 2.37210798169399 | 81.3785750300368 | 90.7174336867795 |
| 75 | Maximum | 1.83 | 80.3626538616887 | 4.14018202793002 | 72.212819395803 | 88.5124883275744 |

Note. Predicted WS and PS values are derived from the combined-device latency models fitted to eligible Eclipse and Chartr observations (n = 298). Robust_SE and the corresponding 95% confidence limits use the HC3 heteroscedasticity-robust covariance estimator; this table contains no p values. Predictions are evaluated at ages 45, 55, 65, and 75 years and at the minimum, first quartile (Q1), median, third quartile (Q3), and maximum observed Wave I latency; remaining continuous covariates are held at their centred value of zero and sex at the model reference setting. Intervals describe uncertainty in the fitted mean score rather than individual-level prediction intervals. WS, word score; PS, phoneme score; SE, standard error; CI, confidence interval.

#### Table S13: Age-stratified WS–PS correlations and HC3-robust regression slopes

| Age_Group | Young (≤median) | Old (>median) | All ages |
|---|---|---|---|
| Overall_Median_Age_Cutoff | 46 | 46 | 46 |
| n | 153 | 145 | 298 |
| Median_Age | 38 | 53 | 46 |
| WS_Mean | 78.4705882352941 | 74.2344827586206 | 76.4093959731543 |
| WS_SD | 5.74348466517246 | 10.0041753735235 | 8.36097510368426 |
| PS_Mean | 91.2712418300653 | 88.56 | 89.9520134228188 |
| PS_SD | 3.13340812439557 | 6.1069223017818 | 4.99495496046343 |
| Correlation_r | 0.885642293565244 | 0.94443055991018 | 0.934126299082537 |
| Correlation_p | 3.63007559628605e-52 | 5.65477712426743e-71 | 1.83844046479248e-134 |
| Slope | 0.48316987329892 | 0.576515688046 | 0.55805916575994 |
| Slope_HC3_SE | 0.0308447835938984 | 0.0363666368717074 | 0.0291015699334574 |
| Slope_HC3_CI_Lower | 0.422226783068699 | 0.50463004067675 | 0.500786963942342 |
| Slope_HC3_CI_Upper | 0.544112963529142 | 0.648401335415249 | 0.615331367577538 |
| Slope_HC3_p | 1.86037233768531e-33 | 2.6514241767171e-33 | 7.75439539192029e-54 |

Note. The age-group cutoff is the overall median age (46 years): younger, age ≤ 46 years; older, age > 46 years. Correlation_r and Correlation_p are the conventional Pearson correlation coefficient and its two-sided p value; these are not HC3 quantities. Slope is from an ordinary least-squares regression of PS on WS. Slope_HC3_SE, Slope_HC3_CI_Lower, Slope_HC3_CI_Upper, and Slope_HC3_p use the HC3 heteroscedasticity-robust covariance estimator. HC3 inference does not constitute adjustment for multiple testing. WS, word score; PS, phoneme score; SD, standard deviation; CI, confidence interval.
